# IL10RB as a key regulator of COVID-19 host susceptibility and severity

**DOI:** 10.1101/2021.05.31.21254851

**Authors:** Georgios Voloudakis, Gabriel Hoffman, Sanan Venkatesh, Kyung Min Lee, Kristina Dobrindt, James M. Vicari, Wen Zhang, Noam D. Beckmann, Shan Jiang, Daisy Hoagland, Jiantao Bian, Lina Gao, André Corvelo, Kelly Cho, Jennifer S. Lee, Sudha K. Iyengar, Shiuh-Wen Luoh, Schahram Akbarian, Robert Striker, Themistocles L. Assimes, Eric E. Schadt, Miriam Merad, Benjamin R. tenOever, Alexander W. Charney, Mount Sinai COVID-19 Biobank, VA Million Veteran Program COVID-19 Science Initiative, Kristen J. Brennand, Julie A. Lynch, John F. Fullard, Panos Roussos

## Abstract

**Background:** Recent efforts have identified genetic loci that are associated with coronavirus disease 2019 (COVID-19) infection rates and disease outcome severity. Translating these genetic findings into druggable genes and readily available compounds that reduce COVID-19 host susceptibility is a critical next step.

**Methods:** We integrate COVID-19 genetic susceptibility variants, multi-tissue genetically regulated gene expression (GReX) and perturbargen signatures to identify candidate genes and compounds that reverse the predicted gene expression dysregulation associated with COVID-19 susceptibility. The top candidate gene is validated by testing both its GReX and observed blood transcriptome association with COVID-19 severity, as well as by *in vitro* perturbation to quantify effects on viral load and molecular pathway dysregulation. We validate the *in silico* drug repositioning analysis by examining whether the top candidate compounds decrease COVID-19 incidence based on epidemiological evidence.

**Results:** We identify *IL10RB* as the top key regulator of COVID-19 host susceptibility. Predicted GReX up-regulation of *IL10RB* and higher *IL10RB* expression in COVID-19 patient blood is associated with worse COVID-19 outcomes. *In vitro* IL10RB overexpression is associated with increased viral load and activation of immune-related molecular pathways. Azathioprine and retinol are prioritized as candidate compounds to reduce the likelihood of testing positive for COVID-19.

**Conclusions:** We establish an integrative data-driven approach for gene target prioritization. We identify and validate *IL10RB* as a suitable molecular target for modulation of COVID-19 host susceptibility. Finally, we provide evidence for a few readily available medications that would warrant further investigation as drug repositioning candidates.

## Introduction

Severe acute respiratory syndrome coronavirus 2 (SARS-CoV-2), which causes coronavirus disease 2019 (COVID-19), is the latest of the betacoronaviruses to pose a global health threat. Of the recent respiratory virus pandemics, SARS-CoV-2 demonstrates the highest transmissibility^1^. Despite the fact that the overwhelming majority of affected individuals have mild symptoms, infection-fatality risk in an urban area of a developed country (e.g. New York City) is still high, ranging from 1.4% for young adults (25-44 years old) to 19.1% for more susceptible older individuals (aged 75 years and older)^2^. Thus, unexplained heterogeneity in susceptibility to the disease and severity of illness exists even when accounting for known risk factors such as age^3^.

The COVID-19 Host Genetics Initiative (HGI)^4^ coordinates a global effort to elucidate the genetic basis of COVID-19 susceptibility. Ongoing efforts have uncovered multiple risk loci for COVID-19 susceptibility; however, these risk variants only partly explain inter-individual variability and, as many of the variants reside within non-coding regions of the genome, the formulation of testable hypotheses to elucidate their potential effects is challenging. To translate these genetic findings to novel therapeutics for COVID-19, we developed a multidisciplinary translational genomics framework that integrates genetic studies of COVID-19 susceptibility, genotype-tissue expression datasets and perturbargen signature libraries to identify druggable gene targets and readily available compounds that can be repositioned as treatments for COVID-19. We provide evidence from *in vitro, in vivo* and retrospective epidemiological studies that validate the association of the top candidate gene, *IL10RB*, with COVID-19 outcome severity. Overall, our study prioritizes gene targets and compounds with direct translational value to modulate host physiology and immune response and increase resilience to SARS-CoV-2 infection.

## Methods

### Transcriptome-wide association study

#### Transcriptomic imputation model construction

Transcriptomic imputation models are constructed as previously described^5^ for peripheral tissues of the GTEx v8^6^ and STARNET^7^ cohorts (Supplementary Table 1; Figure 1A). The genetic datasets of the GTEx and STARNET cohorts are uniformly processed for quality control (QC) steps before genotype imputation. We restrict our analysis to samples with European ancestry as previously described^5^. Genotypes are imputed using the University of Michigan server^8^ with the Haplotype Reference Consortium (HRC) reference panel^9^. Gene expression information is derived from RNA-seq gene level counts which are adjusted for known and hidden confounds, followed by quantile normalization. For GTEx, we use publicly available, quality-controlled, gene expression datasets from the GTEx consortium (http://www.gtexportal.org/). RNA-seq data for STARNET were obtained in the form of residualized gene counts from a previously published study. For the construction of the transcriptomic imputation models we use elastic net based methods; when there is available epigenetic annotation information^10^ for a given tissue we employ the EpiXcan^5^ method to maximize power; when not available, we use the PrediXcan^11^ method. **COVID-19 phenotypes GWAS summary statistics.** Summary statistics for all 7 COVID-19 phenotypes (A1, A2, B1, B2, C1, C2 and D1; Supplementary Table 2) were obtained from the COVID-19 Host Genetics initiative^4^. **Multi-tissue transcriptome-wide association study (TWAS).** We performed the gene-trait association analysis as previously described^5^. Briefly we applied the S-PrediXcan method^12^ to integrate the COVID-19 GWAS summary statistics and the transcriptomic imputation models constructed above to obtain gene-level association results. **Gene set enrichment analysis for TWAS results.** To investigate whether the genes associated with a given trait exhibit enrichment for biological pathways, we use gene sets from MsigDB 5.1^13^ and filter out non-protein coding genes, genes located at MHC as well as genes whose expression cannot be reliably imputed. Statistical significance is evaluated with one-sided Fisher’s exact test and the adjusted p values are obtained by the Benjamini-Hochberg method^14^.

**Figure 1.**
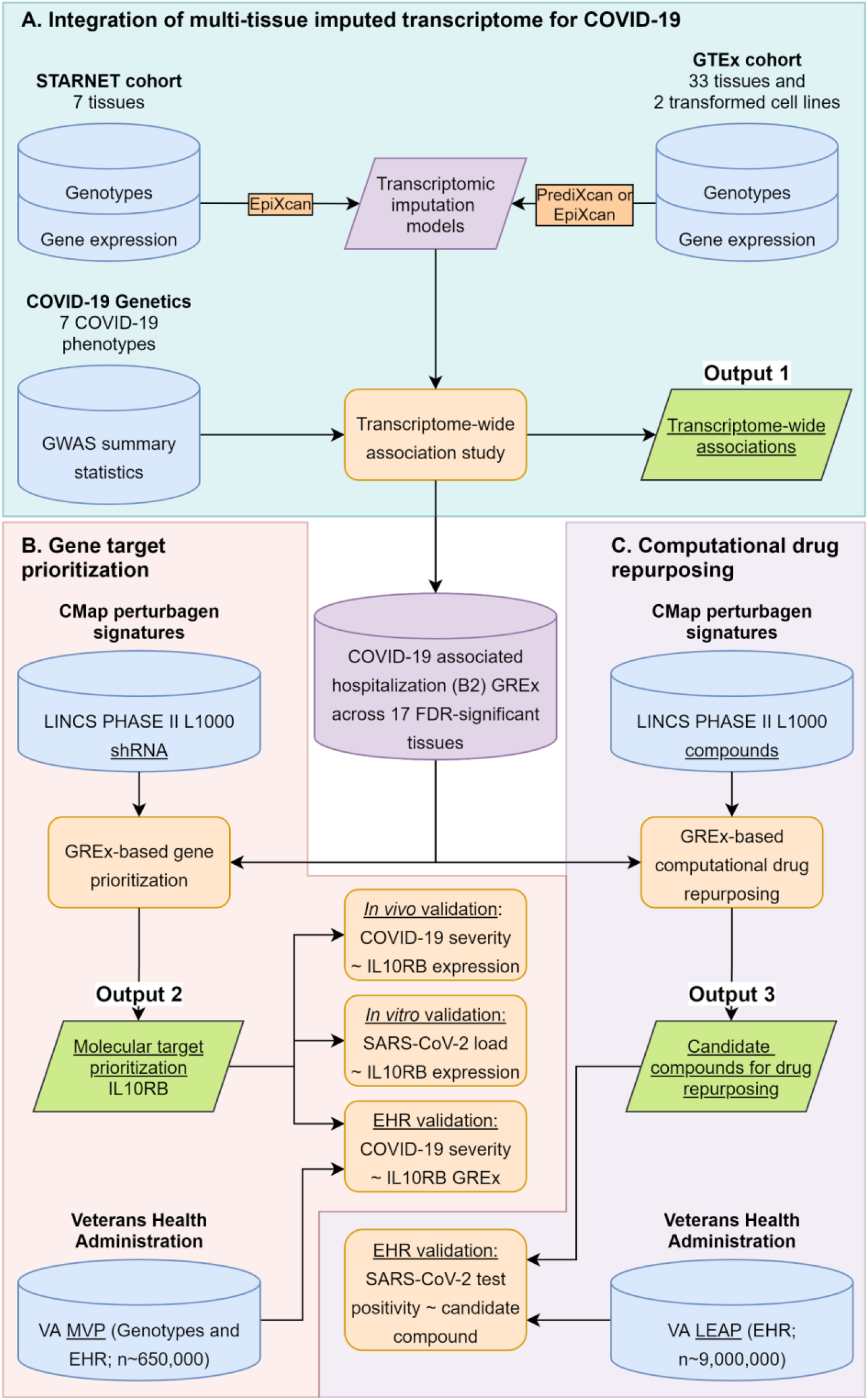
Data-driven GReX-based approach for molecular target prioritization and computational drug repurposing for COVID-19.

### Genetically regulated gene expression (GReX-) based gene targeting approach and computational drug repurposing

Both the gene targeting approach and the computational drug repurposing (CDR) approach integrate genetically regulated gene expression (GReX) information (using the TWAS gene-trait-tissue association results) with a perturbagen signature library^15^ (LINKS Phase II L1000 dataset GSE70138); the gene targeting approach uses the shRNA signatures (gene expression changes after knocking down a gene) and the CDR uses compound signatures (Figure 1). We only consider GReX from 17 tissue models of the B2 phenotype that have significant TWAS results (FDR adjustment is applied to all COVID-19 phenotypes and tissues; Supplementary Table 1; Supplementary Materials and Methods in Supplementary Appendix 1; the steps of the method are also outlined in Supplementary Figure 1). **Signature antagonism of trait GreX.** Each signature from the perturbagen signature library (e.g. *IL10RB* shRNA treatment for 96 hours in MCF7 cells) is assessed and ranked for its ability to reverse the trait-associated imputed transcriptomes using a previously published method^16^. **Summarization of the effect of signatures across tissues.** Signatures are grouped by peturbagen (either shRNA or compound) and we first test whether the signatures for a specific perturbagen are more likely to be ranked higher or lower (Mann-Whitney U test); then we obtain a GReX antagonism pseudo z-score as follows: 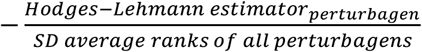. **Gene prioritization approach.** This final step only applies to the gene targeting approach and not the computational drug repurposing. For prioritization we estimate, for each gene, the p value corresponding to the joint statistic of the two approaches (*z*_*combined*=_*<u>z</u>*_*<u>TWAS</u>*_+*pseudo z*_*GreX antagonism*_).

### IL10RB and IFNAR2 GReX association with COVID-19 severity and other phenotypes in the Million Veteran program

#### Cohort

Within the broader cohort of the Million Veteran Program^17^, for the COVID-19 severity analysis we used all COVID19 positive individuals as of March 11^th^, 2021 (n_EUR_=14,262, n_AFR_=5,828, n_HIS_=2,870, n_ASN_=266; EUR: European; AFR: African; HIS: Hispanic; ASN: Asian). For the phenome-wide association study (PheWAS), we used individuals of European Ancestry (n=296,407). Ancestries were defined by the HARE method^18^. Genotypes used for imputation were filtered by Minor Allele Frequency (>0.01), Variant level Missingness (<0.02), as well as imputation R^2^ (>0.9). We consider the MVP severity cohort an independent cohort from the GWAS since less than 7% (1,519) of its participants were included in the COVID-19 related hospitalization GWAS (“B2_ALL_eur_leave_23andme”; release 4) comprising less than 7% and 0.2% of the GWAS’s cases and total individuals respectively. For the individual imputation we use the EpiXcan tissue model of blood from the STARNET cohort for the following reasons: (1) as a tissue, blood is relevant (immune cells) and accessible - allowing for testing and validation, and (2) as an imputation model, the blood (STARNET) is the most powerful model (identifying the most FDR significant gene-trait associations; Supplementary Figure 2), allows the concurrent study of both *IL10RB* and *IFNAR2* (Figure 2a) and is based on collection from beating heart donors (in contrast to the GTEx model which is based mostly on postmortem blood).

**Figure 2.**
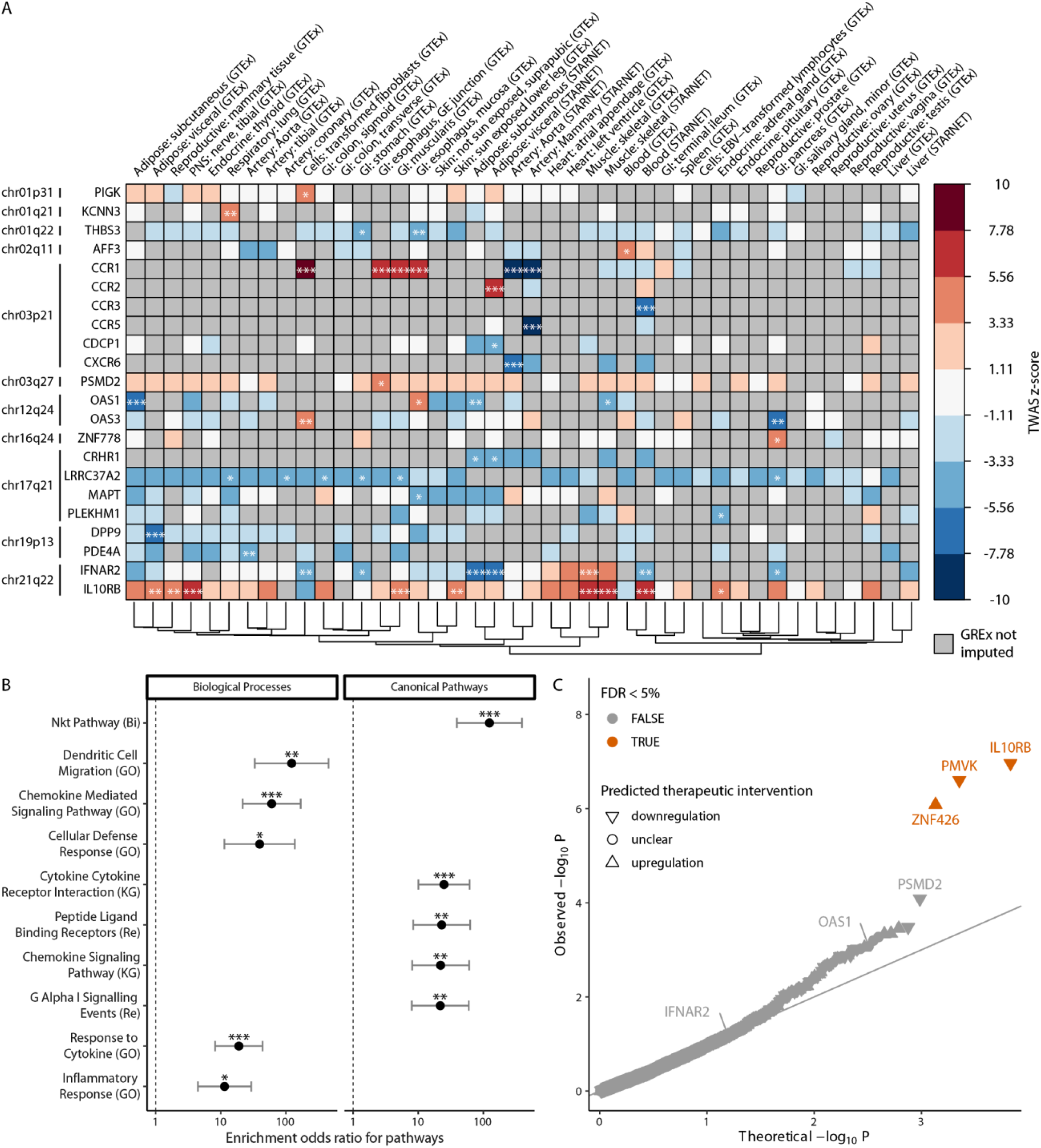
**Transcriptome-Wide Association Study (TWAS) for COVID-19 associated hospitalization (hospitalized COVID vs. the general population) identifies associated genes, pathways and aids in identification of druggable gene targets. Panel A**. FDR-significant TWAS results for COVID-19 susceptibility across all tissues. Box color indicates gene-trait-tissue association z-scores. Gray squares represent genes whose genetically regulated gene expression (GReX) could not be imputed. ***, ** and * correspond to FDR-adjusted p values of association equal or less than 0.001, 0.01 and 0.05 respectively. Dendrogram on the bottom edge is shown from Ward hierarchical clustering for tissues based on all GReX (not just FDR-significant results). Displayed results are limited to protein coding genes; cytogenetic location (at band level resolution) is also provided on the left of each gene. **Panel B**. Enrichment of COVID-19 TWAS associated genes for biological processes and canonical pathways. Odds ratio with 95% confidence interval (CI) is plotted for the significant enrichments of TWAS gene-trait associations from all tissues. Pathways are ranked based on estimated enrichment odds ratio. Analysis is limited to protein coding genes and excludes genes residing in the major histocompatibility complex (MHC) on chromosome 6. Enrichments that are FDR significant are annotated as follows: *, ** and *** for FDR-adjusted p≤0.05, 0.01 and 0.001 respectively; Fisher’s exact test. **Panel C**. Prioritization of candidate gene targets to reverse TWAS gene-trait associations. p value is estimated based on the joint statistic of two approaches (*z*_*combined*=_*<u>z</u>*_*<u>TWAS</u>*+_*pseudo z*_*GReX antagonism*_) against the null. FDR-significant candidate genes are labelled orange. The direction of the predicted therapeutic intervention (upregulation or downregulation) is illustrated. IL10RB, PMVK and ZNF426 are FDR-significant target genes (n=4,302 imputed genes with reliable shRNA signatures).

#### Phenotypes

There are four COVID-19 severity levels: mild, moderate, severe and death (see Supplementary Table 3 for more information of the phenotypic definition and counts).

#### Transcriptomic imputation

GReX for blood *IL10RB* and *IFNAR2* was calculated with the EpiXcan^5^ Blood (STARNET) transcriptomic imputation model (Supplementary Table 1). For TWAS, we only considered SNPs with imputation R^2^≥0.3. Ancestry-specific principal component analysis was performed using the EIGENSOFT^19^ v6 software as previously described^20^.

#### GReX association with COVID-19 severity

Associations of GReX and COVID-19 severity (Supplementary Table 3) were independently performed in each ancestry group. All associations were performed on scaled GReX with the following covariates: Elixhauser comorbidity index^21^ for 2 years, sex, age, age squared (age^2^), and top 10 ancestry-specific principal components. To estimate the effect of GReX on COVID-19 related death we used a logistic regression analysis (binomial distribution) where death was defined as “1” whereas mild, moderate, and severe cases were defined as “0”. To estimate the effect of GReX on COVID-19 related outcome severity we performed an ordinal logistic regression where COVID-19 severity was ordered as follows: mild, moderate, severe and death. The ancestry-specific associations were meta-analyzed with a fixed-effect model using the inverse-variance method to estimate the effect of IL10RB and IFNAR2 GReX in the total population.

#### GReX PheWAS

Phecodes^22^ assigned to clinical encounters up to 2018 (predating the COVID-19 pandemic) were grouped into categories using Phecode Map v1.2 with manual curation for some uncategorized phecodes (as provided in Supplementary Appendix 2). All phecodes with at least one count in more than twenty five individuals in the cohort were considered for further analysis. Association testing was performed between scaled GReX and counts of each phecode with a negative binomial distribution - this is the appropriate distribution to capture the full range of phecode counts since variance was higher than the mean phecode count in 99.95% of the phecodes evaluated (1840/1841; the only exception was “Other disorders of purine and pyrimidine metabolism”). The following covariates were used: total number of phecodes per individual, length of record, sex, age, and top ten ancestry-specific principal components. Phecodes with non-convergent regression models were dropped. Significance was tested at the 0.05 false discovery rate (FDR) level.

### Gene expression profiling and EHR-based phenotyping in the Mount Sinai COVID-19 Biobank

For the Mount Sinai COVID-19 biobank^23^, electronic medical records (EMR) of all patients hospitalized in the previous 24 hours were screened daily to identify eligible patients (and controls) and to obtain bio-specimens with very broad inclusion criteria and very limited exclusion criteria. This study was approved by the Institutional Review Board (IRB) at the Icahn School of Medicine at Mount Sinai (20-0327). Given the extraordinary challenges posed by the COVID-19 pandemic for obtaining informed consent, and in consideration of the public health crisis, a delayed consent model was applied. This delayed consent model enabled bio-specimen collection at the time of clinical blood draws without prior informed consent. Patients were contacted both during their hospital admission, and post-discharge to either provide or decline consent for the use of their banked specimens for research purposes. In the event the patient were to be deemed incapacitated, or expired during their hospitalization, the subject’s legally authorized representative was contacted in order to provide or decline consent. Bio-specimens for this analysis were obtained from 568 individuals; some individuals (n=392) are contributing more than one bio-specimens. The complete biobank dataset and analyses will be presented in Beckmann et al. (manuscript in preparation). In brief, RNA was extracted from blood samples and used to prepare RNA-seq libraries which were QCed and sequenced as previously described^24^. RNA-seq reads were processed, QCed and aligned to a reference genome as previously described^24^. Important covariates affecting transcript counts were identified with the variancePartition method^25^. Cell type proportions were calculated with CIBERSORTx^26^ using the LM22 reference^27^. COVID-19 severity associated cell types are identified as having non-zero coefficients in a linear mixed model lasso procedure (R package glmmlasso) predicting COVID-19 severity. Finally, we used dream^28^ for differential gene expression analysis while accounting for covariates identified by variancePartition^25^ above, as well as, the proportions of COVID-19 severity associated cell types identified above. **COVID-19 severity scale.** Phenotypic information is obtained by the EMR of the Mount Sinai Health System which is reviewed by a screening team that includes practicing physicians. Each bio-specimen is associated, when possible given the information in the EMR, with a COVID-19 severity measurement that corresponds to the time of collection. There are 4 levels of severity^29^: controls, moderate, severe and severe end-organ damage summarized in Supplementary Table 4.

### Manipulation of IL10RB and IFNAR2 expression in cell lines and their effect on SARS-CoV-2 viral load and transcriptional profiles

NGN2-glutamatergic neurons were derived from hiPSC-NPCs of donor NSB2607^30^ as previously described^31^. gRNAs were designed using the CRISPR-ERA (http://crispr-era.stanford.edu) web tool and cloned into a lentiviral transfer vector using Gibson assembly^31,32^; shRNAs were ordered as glycerol stocks from Sigma. Wild type or dCas9 expressing NPCs were infected with rtTA and NGN2 lentiviruses as well as desired shRNA or CRISPRa lentiviruses and differentiated for 7 days before SARS-CoV-2 infection with a multiplicity of infection (MOI) of 0.1 or mock infection for 24 hours. After the completion of the experiment, RNA was isolated, quality metrics were obtained and 200ng of RNA was processed through a total RNA library prep using the KAPA RNA Hyper Prep Kit + RiboErase HMR kit (Roche, cat no: 8098140702) following the manufacturer’s protocol with modifications for automation and optimization to be sequenced on the NovaSeq 6000 S4 2×150bp run at 60M reads per sample. An additional 20ng of RNA was run on a SARS-CoV-2 targeted primer panel using the AmpliSeq Library Plus and cDNA Synthesis for Illumina kits (Illumina, Cat no: 20019103 & 20022654). All samples were normalized, pooled, and run on the NovaSeq 6000 S4 in a 2⨯150 run targeting 750k reads per sample. The STAR aligner v2.5.2a^33^ was used to align reads to the GRCh38 genome (canonical chromosomes only) and Gencode v25 annotation. The module featureCounts^34^ from the Subread package v1.4.3-p1^35^ was used to quantify genes. RSeQC v2.6.1^36^ and Picard v1.77^37^, were used to generate QC metrics. Differential expression analysis was performed with limma^38^ using the first two components of multidimensional scaling and RIN as covariates. Competitive gene set testing using sets from Gene Ontology^39,40^ was performed with camera^41^. SARS-CoV-2 quantification was performed by taxonomically classifying short-read data with taxMaps^42^. The AmpliSeq approach confirmed the presence or absence of the virus in our samples.

### Population-level analysis of the effect of compound and compound category use against COVID-19 incidence

#### Data

We use the VA COVID-19 Shared Data Resource (CSDR), a data domain that includes demographic and clinical information related to COVID-19 of all patients who tested for SARS-CoV-2 within the Veterans Health Administration (VHA) or whose positive test result outside VHA was recorded in VHA clinical notes. The CSDR was supplemented with additional data elements from the Corporate Data Warehouse (CDW) of the VHA, a national repository of national electronic health records of all individuals who received care in the VHA.

#### Cohort

The base cohort included all Veterans who were alive as of February 15, 2020. Since the earliest testing date reported in the CSDR was February 16, 2020, we considered all living patients through February 15 to be eligible to be tested for SARS-CoV-2. From this base cohort, we derived two separate samples to examine incidence of COVID-19 among the users of the top 10 compounds and antiretroviral medications. For the top 10 compound analysis, we assembled a sample consisting of patients who underwent SARS-CoV-2 testing matched to patients who did not undergo SARS-CoV-2 testing on age, race, and VHA facility. The index date was defined as February 15, 2020. For the antiretroviral medication analysis, we created a sample of patients who were ever diagnosed with human immunodeficiency virus (HIV) prior to the index date and were actively on selected antiretroviral medications in the 90 days prior to the index date.

#### Exposure

Exposure to the top 7 FDA-approved compounds (imiquimod, nelfinavir, saquinavir, everolimus, azathioprine, nisoldipine and retinol) and selected antiretroviral medications (see Supplementary Table 5) was determined by collecting prescriptions of the included medications within the 90 days prior to the index date.

#### Outcome

We used a binary variable indicating a positive reverse transcriptase polymerase chain reaction (RT-PCR) SARS-CoV-2 test result through November 30, 2020.

#### Covariates

We assessed at the index date patients’ age, race, marital status, body mass index, smoking status, the Charlson Comorbidity index^43^ in the prior two years, VHA utilization in the prior year, the number of days to first SARS-CoV-2 positivity, and the presence of drug-specific FDA-approved and common off-label indications (Supplementary Table 6; for the individual compound analysis) as determined by International Classification of Diseases. We also included the VHA facility of SARS-CoV-2 testing as a fixed effect in the individual compound models and as a random effect in the antiretroviral medication model.

### Statistical analysis

Multivariable ordinal logistic models were used to test the association of drug exposure with COVID-19 incidence, weighted by the inverse of the predicted probabilities of being tested for SARS-CoV-2. Due to the limited availability of SARS-CoV-2 tests and resources, SARS-CoV-2 testing was prioritized based on a wide ranging factors including patients’ demographics, comorbidities, and severity of symptoms. Such targeted testing likely resulted in a highly selective group of patients who were tested for SARS-CoV-2^44^. To adjust for the non-randomness in testing, we employed the inverse probability weighting method, where the weight is based on the predicted probabilities (propensity scores) of being tested, estimated by a logistic regression model using drug exposure, selected patient covariates, and VHA facility^45,46^. For this propensity model, we implemented the nested case-control design with incidence density sampling to match each tested patient (case) to five patients who were eligible to be tested (controls) at the time of the case’s testing on age, race, and VHA facility^47^.

## Results

### Overview of the multidisciplinary translational genomics framework

We develop a translational genomics framework that integrates three major sources of data (GWAS, genotype-tissue expression datasets and perturbargen signature libraries) to identify and validate susceptibility genes for targeted therapeutics and candidate compounds that are readily available for drug repositioning (Figure 1). We first integrate GWASs for COVID-19 phenotypes with multi-tissue transcriptomic imputation models to predict genetically regulated gene expression (GReX) changes that are associated with COVID-19 susceptibility (Figure 1A; Output 1).

We develop and apply a gene prioritization approach (Figure 1B; Output 2) that integrates GReX with shRNA signature libraries^15^ to identify key genes whose expression: a) is predicted to be dysregulated in COVID-19 susceptible individuals and b) can be targeted to reverse the transcriptome-wide gene expression dysregulation that is associated with COVID-19 susceptibility. From this prioritization step, we identify *IL10RB as* the top gene target candidate, which we subject to three validation steps. The first validation step is to increase phenotypic specificity by examining whether *IL10RB* GReX is associated with COVID-19 severity in patients that tested positive for SARS-CoV-2 (Figure 1B: EHR validation). In addition, we perform a GReX-based PheWAS to further understand the effect of *IL10RB* on pre-existing relevant phenotypes (before the emergence of COVID-19). The second validation step is to associate *IL10RB* gene expression in peripheral blood with COVID-19 severity in a patient cohort (Figure 1B: *in vivo* validation). The third, and final, validation step is to perform isogenic manipulation of *IL10RB* gene expression *in vitro* and study its effect on SARS-CoV-2 viral load and transcriptional dysregulation (Figure 1B: *in vitro* validation).

To identify readily available compounds for drug repositioning, we perform computational drug repurposing and identify top candidates (Figure 1C; Output 3). We then perform population-level analysis to validate these *in silico* predictions by testing whether the candidate compounds decrease the likelihood of testing positive for SARS-CoV-2 (Figure 1C: EHR validation). Overall, this framework provides a prioritized list of: a) novel druggable targets for drug development or repositioning and b) readily available candidate compounds that can be further investigated in clinical trials.

### COVID-19 transcriptome-wide association study

We perform a transcriptome-wide association study (TWAS) leveraging 2 transformed cell lines and 40 peripheral tissue models from two cohorts (GTEx v8^6^ and STARNET^7^; n=16,738 reliably imputed genes; Supplementary Table 1) by using GWAS summary statistics for 7 COVID-19 phenotypes^4^ (Supplementary Table 2). Overall, for COVID-19 phenotypes we observe a very high correlation among the imputed transcriptomes of different tissues (range of Pearson’s *r* is 0.68 to 0.93; Supplementary Figure 3) even though the imputed transcriptomes of the COVID-19 phenotypes are quite diverse (Supplementary Figure 4; more detailed description of the different phenotypes can be found in the Supplementary Appendix 1). 17 genes are significantly associated with COVID-19 infection and outcomes (FDR-adjusted p ≤ 0.05) when considering all 7 phenotypes and 42 tissues: *CCR1, CCR2, CCR3, CCR5, CXCR6, DNPH1, DPP9, IFNAR2, IL10RB, IL10RB-AS1, KCNN3, KIF15, OAS1, OAS2, OAS3, PDE4A* and *TMEM241*. Some of these genes are identified in more than one COVID-19 phenotype (e.g. *IL10RB* and *IFNAR2*) whereas others (e.g. *OAS2*) are only associated with one (Supplementary Figure 5); the GWAS contributing the highest number of gene-trait associations is “hospitalized COVID vs. population” (B2 phenotype as per COVID-19 HGI; 13 out of the 17 genes are captured; Supplementary Figure 5). We also observe that most gene-trait associations are detected in Blood (STARNET) and Mammary artery (STARNET), identifying 7 and 6 gene-trait associations, respectively (Supplementary Figure 2). For the remainder of our analysis we thus focus on the B2 phenotype which we will refer to as “COVID-19 associated hospitalization” as: *(1)* it is the highest powered GWAS and *(2)* conceptually, it captures genetic determinants protecting individuals both from infection (since the control group could have been SARS-CoV-2 negative or non-hospitalized positive) and from severe outcomes of COVID-19 - unsurprisingly, B2 exhibits moderate GReX correlation with both these phenotypes (Supplementary Figure 4).

When considering only the COVID-19 associated hospitalization (B2) phenotype, we identified 88 gene-trait-tissue associations corresponding to 26 unique gene-trait associations (*AFF3, CCR1, CCR2, CCR3, CCR5, CDCP1, CRHR1, CXCR6, DPP9, IFNAR2, IL10RB, IL10RB-AS1, KANSL1-AS1, KCNN3, LINC02210, LRRC37A2, LRRC37A4P, MAPT, OAS1, OAS3, PDE4A, PIGK, PLEKHM1, PSMD2, THBS3, ZNF778*; FDR-adjusted p ≤ 0.05 while only considering B2; Supplementary Appendices 3 to 9 for all results split by COVID-19 phenotype) across 11 genomic regions (Figure 2A for protein-coding genes). Significant genes are enriched for pathways mainly involved in immune host response (Figure 2B). Overall, these results indicate that genetically-associated changes in genes involved in immune related pathways predispose individuals to more severe COVID-19 outcome.

### Gene target prioritization identifies IL10RB as key regulator

Towards prioritizing genes as putative molecular targets for intervention, we next ask the question: “perturbation of which gene would be potentially therapeutic by antagonizing the GReX associated with COVID-19 susceptibility?”. We answer this question with a computational shRNA antagonism approach^5,16^ (Supplementary Figure 1) whose output we integrate with the TWAS findings to identify *IL10RB* as the top significant candidate for gene targeting with a proposed intervention of downregulation (Figure 2C). *IL10RB* is a gene predicted to be significantly upregulated in individuals susceptible to COVID-19 hospitalization and whose downregulation significantly antagonizes the polygenetically driven gene expression differences (GReX) associated with COVID-19 hospitalization (Supplementary Figure 2C left panel). Based on mouse models, IL10RB overexpression increases susceptibility to lethal bacterial superinfections in the lung both via postviral increased IL-10 signaling which dampens the immune response^48^, and by direct disruption of the lung epithelial barrier via increased expression of type III interferons (IFNλ)^49^. *IFNAR2*, which is in the same locus (less than 2kbp from *IL10RB*), is not significant in this analysis, thereby nominating *IL10RB* as the most promising candidate in the locus, while deprioritizing *IFNAR2*.

### Predicted upregulation of blood IL10RB is associated with COVID-19 severity and increased incidence of respiratory failure

The COVID-19 related hospitalization GWAS has utilized a broadly-defined phenotype to increase cohort inclusion and sample size. To enhance granularity and phenotyping depth of IL10RB GReX association with COVID-19 associated hospitalization, we determine whether predicted up-regulation of blood IL10RB predicts COVID-19 outcome severity and death in individuals who tested positive for SARS-CoV-2. We perform individual GReX imputation and association analysis in the VA’s Million Veteran Program (MVP)^17^, where severity of COVID-19 related outcomes is deduced from EHR of COVID-19 positive cases (n=23,226; cohort characteristics in Supplementary Table 7). *IL10RB* GReX is associated with increased incidence of COVID-19 related death in individuals of European descent (EUR; logistic regression; OR=1.13; Bonferroni-adjusted p=0.01; n=14,262) and in the trans-ethnic meta-analysis (logistic regression; OR=1.12; Bonferroni-adjusted p=0.002; n=23,226) (Figure 3A and Supplementary Figure 6); *IFNAR2* GReX is not associated with COVID-19 death. However, both *IL10RB* and *IFNAR2* GReX are associated with more severe COVID-19 clinical outcomes in EUR, participants of African descent (AFR) and in the transethnic meta-analyses (ordinal logistic regression; Figure 3A and Supplementary Figure 6; Supplementary Table 8).

**Figure 3.**
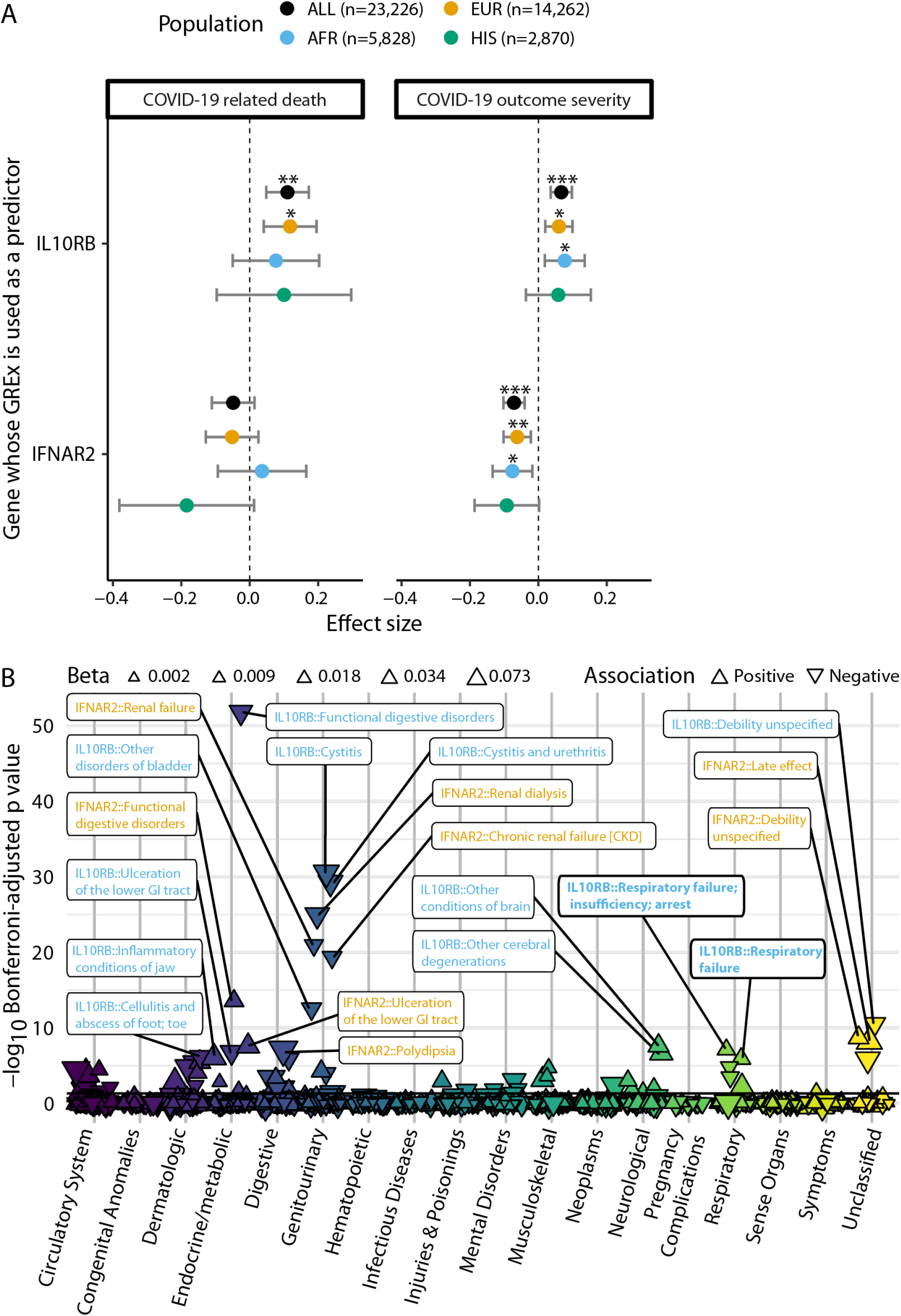
Association of blood IL10RB and IFNAR2 GReX with COVID-19 related outcomes and non COVID-19 phenotypes. Panel A. GReX of IL10RB and IFNAR2 is imputed in 23,216 individuals in the MVP cohort for whom COVID-19 outcome severity information is available. For COVID-19 related death (left panel) we check the association of GReX with the outcome of COVID-19 related death (4.8% of this cohort) under logistic regression models for *IL10RB* and *IFNAR2* GReX while adjusting for age, sex, Elixhauser’s comorbidity score and ancestry-specific population structure. For COVID-19 outcome severity, we applied an ordinal regression model (same predictors and covariates as above) using an outcome scale corresponding to mild (74.9% of the cohort), moderate (17%), severe (3.2%) COVID-19 related outcomes and death (4.8%). EUR, AFR and HIS refer to harmonized European, African and Hispanic ancestry respectively and the sample sizes are provided in the legend at the top. For both panels, the population of Asian ancestry (n = 266) is included in the fixed effects meta-analysis (Population: “ALL” in the graph) but not plotted. ***, ** and * correspond to Bonferroni-adjusted association p values (for n_genes_ × n_outcomes_ for each population cohort) of equal or less than 0.001, 0.01 and 0.05 respectively. **Panel B**. PheWAS of IL10RB and IFNAR2 blood GReX for individuals of European descent in the MVP cohort (n=296,407). Phenotypes are grouped in categories shown in the x-axis, y-axis represents −log_10_(Bonferroni-adjusted p values). The data points are triangles that face to the top or bottom for positive and negative association with GReX respectively; the magnitude of the effect size is represented by the triangle size and the color corresponds to the phenotype category. Only the top 20 associations are labeled (orange for IFNAR2 and blue for IL10RB); full results are provided in Supplementary Appendix 10. Horizontal black line corresponds to Bonferroni-adjusted p = 0.05.

To better understand the phenotypic variation linked with *IL10RB* and *IFNAR2* imputed expression, we perform a phenome-wide association study (PheWAS) utilizing the *IL10RB* and *IFNAR2* GReX models in MVP (Figure 3B; cohort characteristics in Supplementary Table 7; Supplementary Appendix 10 for complete set of results). For *IL10RB*, among significant results, we found that COVID-19 related GReX dysregulation (higher *IL10RB* GReX in blood) was positively associated with respiratory failure and tracheostomy complications and disorders of the circulatory system such as heart aneurysms and non-rheumatic mitral valve disorders as well as cholecystitis without cholelithiasis and inflammatory conditions of the jaw. On the other hand, it was negatively associated with intracerebral hemorrhage, infections of the skin (e.g. lower limb cellulitis) and genitourinary system (e.g. cystitis and urethritis), type 1 diabetes, kidney disease (e.g. renal osteodystrophy), schizophrenia, functional disorders of the digestive system and bladder, and overall unspecified debility and sequela. COVID-19 related *IFNAR2* GReX dysregulation (lower *IFNAR2* GReX in blood) shares some positive associations with *IL10RB* such as respiratory failure and heart aneurysms but is independently associated with congestive heart failure, (chronic) renal failure and dialysis, delirium dementia, stomach cancer and antisocial/borderline personality disorder. On the other, we identify negative associations with cerebral ischemia, specific infections (cellulitis of foot, toe and pyoderma, acute osteomyelitis), arthropathies, functional disorders of the digestive system and bladder, and overall unspecified debility and sequela. Overall, in addition to predisposing individuals to COVID-19 related hospitalization and outcome severity, increased *IL10RB* and decreased *IFNAR2* GReX are associated with respiratory failure independent of COVID-19 exposure.

### Increased *IL10RB* blood expression predicts worse COVID-19 outcome

Transcriptome imputation models can only partially explain the variance in observed *IL10RB* and *IFNAR2* gene expression (R^2^_CV_ is 0.099 and 0.278 respectively). To further confirm the association of IL10RB with COVID-19 severity, we utilize blood gene expression profiling data from COVID-19 patients and controls at the Mount Sinai COVID-19 Biobank^23^ (cohort characteristics in Supplementary Table 9 and Supplementary Table 10). We establish a direct significant association between observed blood *IL10RB* expression and severe COVID-19 outcome, including end-organ damage. The levels of *IL10RB* expression are gradually increased with disease severity with higher effect size in the most severe COVID-19 patient group (end-organ damage) against all other groups (Figure 4A). Similar analysis for blood *IFNAR2* gene expression failed to demonstrate a robust association.

**Figure 4.**
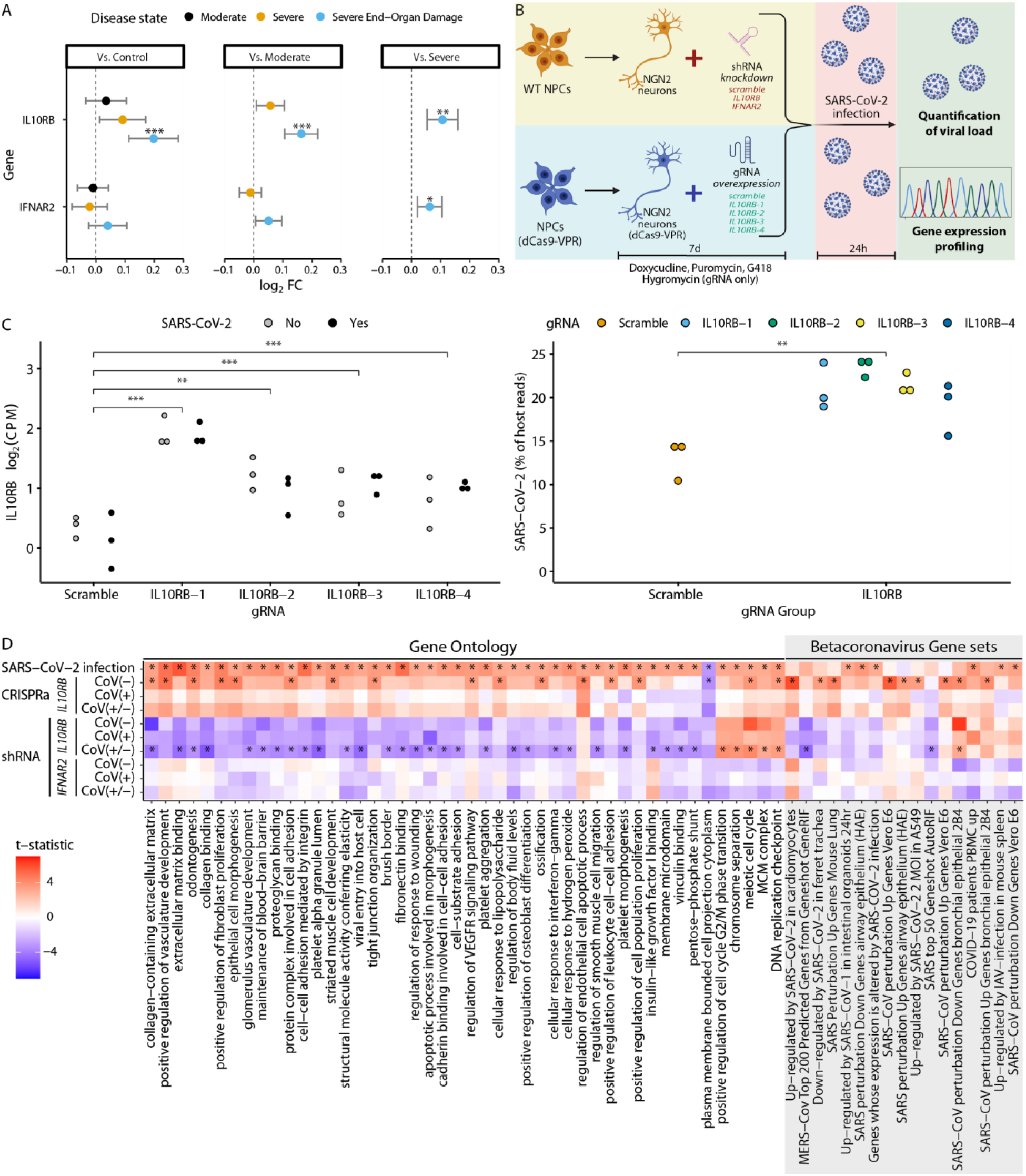
Increased IL10RB expression is associated with worse COVID-19 outcomes *in vivo* and increased SARS-CoV-2 viral load *in vitro*. **Panel A**. Increased IL10RB expression is associated with worse COVID-19 outcomes *in vivo*. *, ** and *** for FDR-adjusted p≤0.05, 0.01 and 0.001 respectively **Panel B**. *In vitro* experimental overview. **Panel C**. CRISPRa gRNAs (IL10RB-1, IL10RB-2, IL10RB-3 and IL10RB-4) were used to overexpress IL10RB in hiPSC-derived NGN2-glutamatergic neurons. ***, ** and * correspond to p values from the linear model of equal or less than 0.001, 0.01 and 0.05 respectively. For the SARS-CoV-2 viral load (right panel) we perform pairwise comparison with unpaired t-test; ***, ** and * correspond to p values equal to, or less than, 0.001, 0.01 and 0.05 respectively. **Panel D**. Competitive gene set enrichment analysis in hiPSC-derived NGN-2 glutamatergic neurons. On the left side we are testing enrichment for canonical pathways and biological processes (Gene ontology) that are significantly enriched (FDR<0.05) in SARS-CoV-2 infection (top row) and on the right side for gene sets that correspond to betacoronavirus infections across different cell systems^52^ (only significant results are shown; FDR < 0.05).

### *IL10RB* overexpression increases *in vitro* SARS-CoV-2 viral load

It has recently been shown that SARS-CoV-2 viral load in patients is associated with increased disease severity and mortality^50^. To explore the effect of *IL10RB* and *IFNAR2* expression on SARS-CoV-2 viral load we perform a series of *in vitro* experiments where we manipulate gene expression levels with short hairpin (shRNA; for down-regulation) and clustered regularly interspaced short palindromic repeats activation (CRISPRa; for up-regulation) and quantify the SARS-CoV-2 viral load (Figure 4B). Overall, we test (in technical triplicates) down-regulation of *IL10RB* and *IFNAR2* by shRNA (Supplementary Figure 7; Supplementary Figure 8) and up-regulation of *IL10RB* by CRISPRa (by using four different guide RNAs; Figure 4C). We perform these experiments in NGN2-glutamatergic post-mitotic neurons^32^ derived from human induced pluripotent stem cell (hiPSC) since these cells can be effectively infected by SARS-CoV-2, allow replication of the virus and can serve as a model cell system for SARS-CoV-2^51^. Indeed, we find that the gene expression dysregulation caused by SARS-CoV-2 infection in our model cell system mimics the transcriptional signatures corresponding to SARS-CoV-2 (and other betacoronaviruses) infection of a diverse range of cell types^52^ (Figure 4D; Supplementary Figure 9).

We observe a significant increase in SARS-CoV-2 viral load (Figure 4C; p=0.0087; unpaired t-test) after IL10RB overexpression using 4 different guide RNAs (gRNAs) (Figure 4C and Supplementary Table 11). Competitive pathway enrichment analysis demonstrates that overexpressing *IL10RB* in non-infected cells leads to the induction of COVID-19 relevant pathways implicated in vascular, immune system and extracellular matrix processes (Figure 4D), which are also activated by SARS-CoV-2 infection (Figure 4D). Surprisingly, even in the absence of SARS-CoV-2, *IL10RB* over-expression leads to transcriptional changes reminiscent of betacoronavirus infection (Supplementary Figure 9). In the rescue experiment, shRNA knock-down of *IL10RB* does not reduce *IL10RB* levels robustly, most likely due to low basal expression (Supplementary Figure 7; Supplementary Table 12. On the other hand, we are able to successfully knock down *IFNAR2* (higher basal expression; Supplementary Figure 8; Supplementary Table 12) which leads to a decrease in SARS-CoV-2 load. Interestingly, SARS-CoV-2 infection induces expression of *IFNAR2* (Supplementary Figure 8; Supplementary Table 13) but not *IL10RB* (Supplementary Figure 7; Supplementary Table 13). This suggests that the increased *IFNAR2* (but not *IL10RB*) levels observed in the most severe group of COVID-19 patients (Figure 4A) may reflect the increased likelihood of SARS-CoV-2 viremia in those patients^50^.

### Computational drug repurposing analysis and population-level validation of top candidate compounds against COVID-19 incidence

We perform computational drug repurposing to identify compounds^53^ with the potential to reverse the GReX associated with COVID-19 related hospitalization. The top 10 candidates are imiquimod, nelfinavir, saquinavir, everolimus, azathioprine, nisoldipine, cerulenin, pyrvinium-pamoate, retinol and selamectin (Supplementary Table 5; all results in Supplementary Appendix 11). After excluding the compounds that are not currently FDA approved (cerulenin, pyrvinium-pamoate and selamectin), we determine whether a compound is associated with reduced COVID-19 incidence. We identify 755,346 veterans who have received the SARS-CoV-2 test from the broader Veterans Health Administration cohort (over 9 million US veterans) and estimate the likelihood of a positive SARS-CoV-2 test when the candidate compound is prescribed within 90 days prior to the test. We exclude from the analysis nelfinavir, saquinavir and nisoldipine (Supplementary Table 14), as they have been prescribed in less than 100 individuals. For the final population-level analysis, we consider the remaining four compounds: imiquimod, everolimus, azathioprine and retinol. We also group compounds by mechanism of action, no mechanism of action was significant when adjusting for multiple test correction but the top class was anti-HIV protease inhibitors (Supplementary Table 5).

In the individual compound analysis (Supplementary Appendix 12), 2 out of 4 compounds are significantly associated with a lower likelihood of testing positive for SARS-Cov-2 (Figure 5) after adjusting for important epidemiologic factors, medication indications and propensity to get tested (Supplementary Table 6; Supplementary Appendix 12). Azathioprine (odds ratio, 0.69; 95% CI, 0.62 to 0.77) and retinol (oral administration; odds ratio, 0.81; 95% CI, 0.72 to 0.92) are significantly associated with a strong decrease in COVID-19 incidence (Figure 5). Imiquimod, which is administered in the form of a topically applied cream (either 3.75% or 5%), has been shown to have significant systemic absorption both in humans^54^ and mouse models^55^ but it is unclear if it is sufficient to have a systemic effect. For everolimus we had an order of magnitude fewer exposed individuals so we may not be sufficiently powered to detect an effect. When grouping medications by mechanism of action, the top category is anti-HIV protease inhibitors. Given how HIV medications are currently prescribed in highly active antiretroviral therapy (HAART) cocktails, we perform the analysis within HIV patients receiving antiretroviral treatment and compare the individuals who take anti-HIV protease inhibitors against the individuals who take other antiretroviral medications while adjusting for important covariates, as with the individual medication analysis above; overall these two groups have similar medical comorbidities (Supplementary Appendix 12). We find no effect of anti-HIV protease inhibitors against COVID-19 incidence (Figure 5; Supplementary Appendix 12).

**Figure 5.**
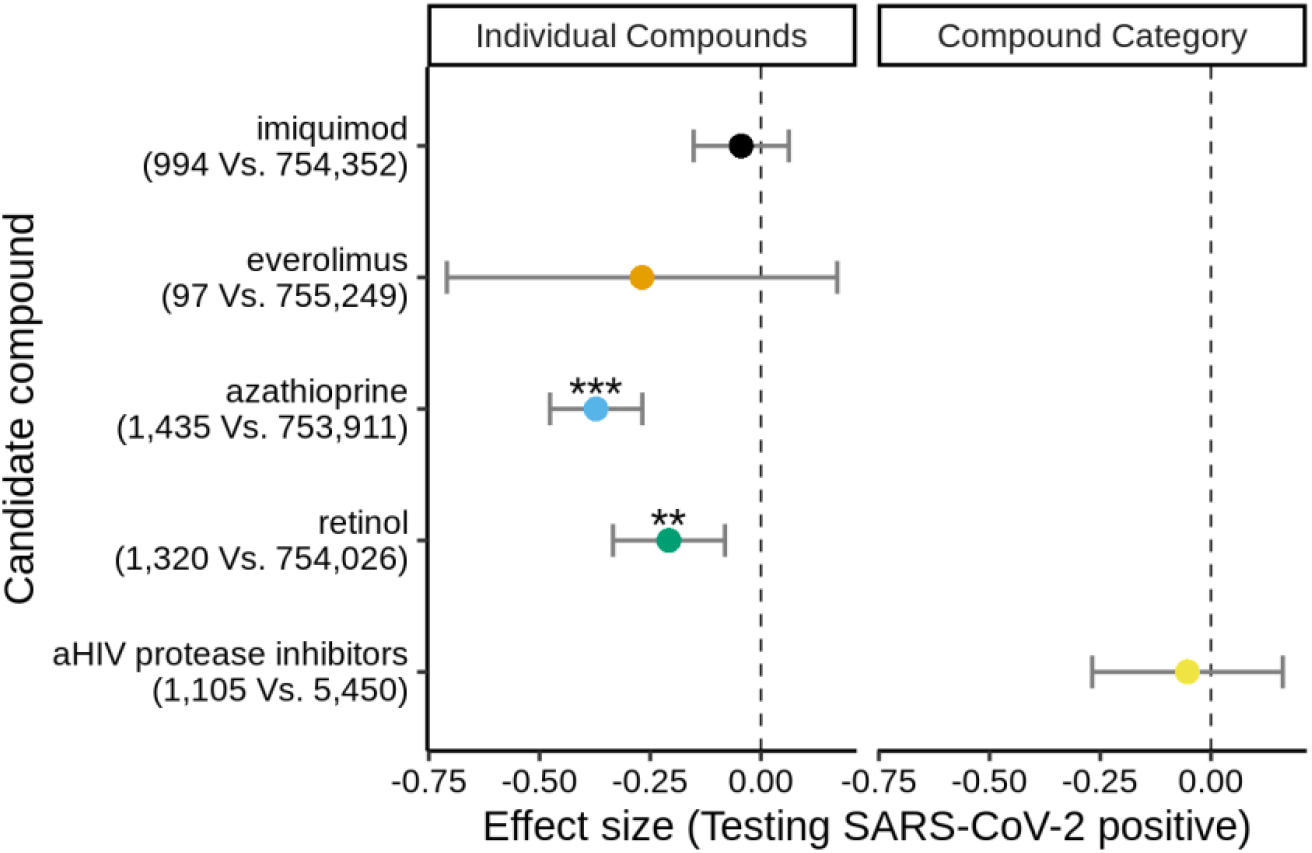
EHR validation of computational drug repurposing. Top individual compounds (imiquimod, everolimus, azathioprine and retinol; left panel) and the top compound category (anti-HIV protease inhibitors; right panel) are evaluated to determine whether they are associated with a decreased likelihood for a positive SARS-CoV-2 test. For the individual compound analysis, all individuals that had a SARS-CoV-2 test and could be matched are considered (n=755,346). The number of individuals that tested positive vs. negative for SARS-CoV-2 are provided in the y-axis labels. For the top compound category (anti-HIV protease inhibitors), the group of those receiving anti-retroviral treatment with an anti-HIV protease inhibitor (n=1,105) are compared against those that are receiving other anti-HIV medications (n=5,450). The effect size (log(OR)) is plotted along with 95% CI error bars. ***, ** and * correspond to Bonferroni-adjusted association p values equal to, or less than, 0.001, 0.01 and 0.05, respectively.

## Discussion

Our multidisciplinary translational genomics framework integrates genetic variation, GREx and perturbargen signature libraries to identify druggable gene targets and readily available compounds that can be repositioned for COVID-19 (Figure 1). Transcriptomic imputation^5,11,12^ serves as the genomics backbone of this approach and it trades off a part of SNP heritability in exchange for GReX^56^ which has translational potential. For gene target prioritization, this potential is realized with the integration of the multi-tissue TWAS results with an shRNA signature library^15^ to identify genes whose perturbation can reverse the disease-associated GReX. This shRNA GReX antagonism approach identifies *IL10RB* (21q22.11) as the most promising gene target and overcomes traditional limitations of GWAS and TWAS analyses in identifying key genes within a gene cluster. Based on existing approaches, *IL10RB* would not have been the top candidate for further investigation. First, the index SNP for COVID-19 susceptibility in 21q22.11, rs13050728^57^, falls within an intronic region of *IFNAR2* (less than 24kbp from *IL10RB*). In addition, integrating genotype-gene expression datasets cannot identify the most likely causal gene in this locus since the index SNP is associated with gene expression changes of both *IFNAR2 and IL10RB*^*57*^. Similarly, the genes can only be partially prioritized with targeted individual imputation (Figure 3A; *IFNAR2* is not associated with COVID-19 death but is associated with COVID-19 severity) and cannot be prioritized on TWAS based on summary statistics (Figure 2A) even when considering splicing^58^ due to co-regulation^59^ (Supplementary Figure 10). *IL10RB* is, in part, prioritized because it has a more uniform imputed transcriptional dysregulation across tissues (predominant down-regulation; Figure 2A, Supplementary Figure 11). On the other hand, *IFNAR2* is expected to be up-regulated in some tissues and down-regulated in others^58^ (Figure 2A, Supplementary Figure 11), e.g. there is a consistent, predicted, down-regulation in adipose tissue and an opposing up-regulation in muscle tissue (Supplementary Figure 12). Unfortunately, technological innovations that would allow differential targeting of tissues are not readily available; thus, our gene target prioritization approach inherently penalizes opposing effects on GReX by integrating multiple tissues (Supplementary Figure 1).

Towards validating *IL10RB* as a suitable molecular target, we establish a direct association of increased *IL10RB* GReX (Figure 3A) and expression (Figure 4A) with worse COVID-19 clinical outcomes and death. Importantly, these results provide external validity for our findings beyond individuals of European ancestry^60^ by performing ancestry-specific analysis for GReX (Figure 3A) and leveraging a diverse patient cohort for expression profiling (Figure 4A). Finally, isogenic manipulation of *IL10RB* in a model cell system for SARS-CoV-2^51^ reveals that inducing *IL10RB* expression results in priming of SARS-CoV-2 pathways (Figure 4D) and increased SARS-CoV-2 viral load upon infection (Figure 4C) which is associated with worse COVID-19 outcomes^50^. IL10RB serves as a receptor for members of the extended IL-10 family of cytokines (IL10RA_2_IL10RB_2_ heterotetramer for IL-10; IL10RB heterodimers with IL22RA1, IL20RA and IFNLR1 for IL-22, IL-26 and IFNL1/IFNL2/IFNL3 respectively) which emerged before the adaptive immune response and are essential in modulating host defense mechanisms, especially in epithelial cells, to limit the damage caused by viral and bacterial infections^61^. This family of ligands has diverse and often contradicting roles in host response with an undetermined extent of functional cross-talk between them^62^; thus, further molecular dissection will be required to identify the causal signaling pathway(s) of IL10RB in COVID-19 susceptibility. IL-10 was found to be an important mediator of enhanced susceptibility to respiratory post-viral bacterial superinfections in a mouse model^48^. IL-22 promotes antibacterial activity^63^ and enhances tissue regeneration and wound healing^64^. IL-26 is poorly understood. Finally, IFN-λs (IL-28/IL29) are induced by viral infection and show antiviral activity^65,66^; unsurprisingly, in a COVID-19 mouse model administration of IFN-λ1a diminishes SARS-CoV-2 replication^67^. However, the participation of IFN-λs in damaging pro-inflammatory responses remains to be evaluated since recent mouse model studies showed that: 1) viral RNA induced IFN-λ production causes direct disruption of the lung epithelial barrier and increases susceptibility to bacterial superinfections^49^ and 2) IFN-λ signaling aggravates viral infection by impairing lung epithelial regeneration^68^. Clinical outcomes from pegylated IFN-λ1a clinical trials against COVID-19 will provide evidence about the desired modulation direction of this pathway in COVID-19 treatment (phase II clinical trials: NCT04343976, NCT04354259, NCT04534673 and NCT04344600). Possible next steps include the evaluation of readily available IL-10 neutralizing antibodies (e.g. BT063, Biotest; NCT02554019) and IL-22 neutralizing antibodies (e.g. ILV-094/095, Pfizer). While the molecular dissection of this COVID-19 susceptibility pathway is important, transiently down-regulating *IL10RB* with RNA interference (RNAi^69^) may be sufficient if a lung targeting approach is developed.

We identify currently available medications that have the potential to reverse the polygenic nature of gene expression dysregulation associated with COVID-19 related hospitalization by applying a GReX-based computational drug repurposing approach^5,16^ in a separate analytic arm. We previously performed a validation of the overall approach and showed that compounds that were predicted to be therapeutic exhibited a progressive enrichment for higher physician-curated indication levels across a wide range of diseases (e.g. cardiovascular, autoimmune and neuropsychiatric)^5^. In this study, since there are no known treatments for COVID-19, we validate our approach by performing a population-level analysis to study the effect of candidate compounds against COVID-19 susceptibility. One of the limitations of this study is that, due to limited power, we are studying the effect of the compounds on COVID-19 incidence rather than COVID-19 severity. Since we have no direct evidence to support an effect of the compounds on COVID-19 disease progression, we can only provide recommendation for prioritization of compounds for follow-up *in vitro* and *in vivo* studies. The only compounds identified by the *in silico* analysis which performed well in the retrospective epidemiologic validation (Figure 5) are azathioprine and retinol. Immunosuppressive therapy (including azathioprine as a monotherapy or combination therapy) has been proposed to be beneficial in a small retrospective study^70^ but there is conflicting evidence regarding azathioprine treatment in a COVID-19 ferret model^71^. For retinol (vitamin A), a recent study of molecular simulations identified it as a potential ligand that may stabilize the closed conformation of the spike protein thus possibly reducing the opportunity for ACE2 interaction^72^; alternative mechanisms have been hypothesized but not experimentally tested^73–76^. In pursuit of a candidate medication class, the negative findings of the anti-HIV protease inhibitor population-level analysis (Figure 5) echo the recent results of the randomized controlled clinical trial that found no benefit in prescribing lopinavir-ritonavir in hospitalized adult patients^77^. Our computational drug repurposing pipeline positioned anti-HIV protease inhibitors in the first place due to nelfinavir and saquinavir (Supplementary Table 5) but these medications were prescribed to less than 0.5% of the individuals receiving anti-HIV protease inhibitors in the validation cohort (Supplementary Table 15). Thus, even though we have demonstrated that there is no significant effect of the anti-HIV protease inhibitors as a medication class, nelfinavir and saquinavir may still be valuable candidates. In particular, nelfinavir is a promising compound worthy of further exploration based on a recent study highlighting its excelling anti-SARS-CoV-2 *in vitro* activity and pharmacokinetic profile among the anti-HIV protease inhibitors that were tested^78^. In addition to the high anti-SARS-CoV-2 activity (EC_50_=1.13μM), selectivity index (CC_50_/EC_50_=23.32), C_trough_/EC_50_ ratio (3.43)^78^ and potent inhibition of SARS-CoV-2 spike (S) glycoprotein mediated cell fusion at ∼10μM that would prevent the virus to avoid extracellular neutralizing antibodies^79^, we now provide GReX-based evidence for nelfinavir to antagonize predicted gene expression dysregulation in susceptible hosts.

## Data Availability

All data will be made available

## Funding and Disclosures

This research is based on data from the Million Veteran Program, Office of Research and Development, Veterans Health Administration, and was supported by award #MVP035. This publication does not represent the views of the Department of Veteran Affairs or the United States Government. This study was also supported by the National Institutes of Health (NIH), Bethesda, MD under award numbers K08MH122911 (Georgios Voloudakis), R01AG050986, R01AG065582, R01AG067025, U01MH116442, R01MH109677 (Panos Roussos), and by the Veterans Affairs Merit grants BX002395 and BX004189 (Panos Roussos). This study has also been funded in part by the Brain & Behavior Research Foundation via the 2020 NARSAD Young Investigator Grant #29350 (Georgios Voloudakis) and the Icahn School of Medicine at Mount Sinai via a COVID-19 Pilot Grant (Schahram Akbarian, Benjamin R. tenOever, Kristen J. Brennand).

## Notes

### Competing Interest Statement

The authors have declared no competing interest.

### Author Declarations

GReX analysis in MVP: These analyses were conducted under the protocol VA Million Veteran Program COVID-19 Science Initiative, which was approved by the Veterans Affairs Central Institutional Review Board and by the Research & Development Committee at the James J. Peters VA Medical Center.

Gene expression profiling and EHR-based phenotyping in the Mount Sinai COVID-19 Biobank: This study was approved by the Institutional Review Board (IRB) at the Icahn School of Medicine at Mount Sinai (20-0327).

Population-level analysis of the effect of compound and compound category use against COVID-19 incidence: Analysis of national VA data was conducted under the protocol, Leveraging Electronic Health Information to Advance Precision Medicine (LEAP), which was approved by VA Central Institutional Review Board and by the Research & Development Committees at Palo Alto, Salt Lake City, and West Haven VA Medical Centers.

