## Supplementary Appendix 1 for "IL10RB as a key regulator of COVID-19 host susceptibility and severity"

**SUPPLEMENTARY NOTE** 3
VA Million Veteran Program COVID-19 Science Initiative: Core Acknowledgment for Publications 4
MVP COVID-19 Science Program Steering Committee 4
MVP COVID-19 Science Program Steering Committee Support 4
MVP COVID-19 Science Program Working Groups and Associated Chairs 4
MVP COVID-19 Genomics and PRS Working Group Roster 6
MVP COVID-19 GWAS Working Group Roster 7
MVP Executive Committee 8
MVP Program Office 8
MVP Recruitment/Enrollment 9
MVP Science 9
Current MVP Local Site Investigators 10
Mount Sinai COVID-19 Biobank: Core Acknowledgment for Publications 14
Other Acknowledgements 15

**SUPPLEMENTARY MATERIALS AND METHODS** 16
Transcriptome-wide association study 17
Genetically regulated gene expression (GReX-) based gene targeting approach and computational drug repurposing 17
Gene expression profiling and EHR-based phenotyping in the Mount Sinai COVID-19 Biobank (full methods, dataset and results will be presented in Beckmann et al, manuscript in preparation). 18
Manipulation of IL10RB and IFNAR2 expression in cell lines and their effect on SARS-CoV-2 viral load and transcriptional profiles 18

**SUPPLEMENTARY RESULTS** 22
COVID-19 phenotypes genetically regulated gene expression (GREx) comparison. 22

**SUPPLEMENTARY FIGURES** 23
Supplementary Figure 1 24
Supplementary Figure 2 26
Supplementary Figure 3 27
Supplementary Figure 4 28
Supplementary Figure 5 29
Supplementary Figure 6 30
Supplementary Figure 7 31
Supplementary Figure 8 32
Supplementary Figure 9 33

Supplementary Figure 10 34
Supplementary Figure 11 35
Supplementary Figure 12 36

**SUPPLEMENTARY TABLES 37**
Supplementary Table 1 38
Supplementary Table 2 40
Supplementary Table 3 41
Supplementary Table 4 42
Supplementary Table 5 43
Supplementary Table 6 44
Supplementary Table 7 47
Supplementary Table 8 48
Supplementary Table 9 49
Supplementary Table 10 50
Supplementary Table 11 51
Supplementary Table 12 52
Supplementary Table 13 53
Supplementary Table 14 54
Supplementary Table 15 55
Supplementary Table 16 56

**DESCRIPTION OF OTHER SUPPLEMENTARY APPENDICES 57**
SUPPLEMENTARY APPENDIX 2 58
SUPPLEMENTARY APPENDICES 3 TO 9 58
SUPPLEMENTARY APPENDIX 10 60
SUPPLEMENTARY APPENDIX 11 61
SUPPLEMENTARY APPENDIX 12 61

**REFERENCES 62**

### SUPPLEMENTARY NOTE

### VA Million Veteran Program COVID-19 Science Initiative: Core Acknowledgment for Publications

#### MVP COVID-19 Science Program Steering Committee


- Christopher J. O'Donnell, M.D., M.P.H. (Co-Chair)
  VA Boston Healthcare System, 150 S. Huntington Avenue, Boston, MA 02130
- J. Michael Gaziano, M.D., M.P.H. (Co-Chair)
  VA Boston Healthcare System, 150 S. Huntington Avenue, Boston, MA 02130
- Philip S. Tsao, Ph.D. (Co-Chair)
  VA Palo Alto Health Care System, 3801 Miranda Avenue, Palo Alto, CA 94304
- Sumitra Muralidhar, Ph.D.
  US Department of Veterans Affairs, 810 Vermont Avenue NW, Washington, DC 20420
- Jean Beckham, Ph.D.
  Durham VA Medical Center, 508 Fulton Street, Durham, NC 27705
- Kyong-Mi Chang, M.D.
  Philadelphia VA Medical Center, 3900 Woodland Avenue, Philadelphia, PA 19104
- Juan P. Casas, M.D., Ph.D.
  VA Boston Healthcare System, 150 S. Huntington Avenue, Boston, MA 02130
- Kelly Cho, M.P.H., Ph.D.
  VA Boston Healthcare System, 150 S. Huntington Avenue, Boston, MA 02130
- Saiju Pyarajan, Ph.D.
  VA Boston Healthcare System, 150 S. Huntington Avenue, Boston, MA 02130
- Jennifer Huffman, Ph.D.
  VA Boston Healthcare System, 150 S. Huntington Avenue, Boston, MA 02130
- Jennifer Moser, Ph.D.
  US Department of Veterans Affairs, 810 Vermont Avenue NW, Washington, DC 20420


#### MVP COVID-19 Science Program Steering Committee Support


- Lauren Thomann, M.P.H. (P&P Committee Representative, Working Group Coordinator)
- VA Boston Healthcare System, 150 S. Huntington Avenue, Boston, MA 02130
- Helene Garcon, M.D. (Program Coordinator, Working Group Coordinator)
  VA Boston Healthcare System, 150 S. Huntington Avenue, Boston, MA 02130
- Nicole Kosik, M.P.H. (Working Group Coordinator)
  VA Boston Healthcare System, 150 S. Huntington Avenue, Boston, MA 02130


#### MVP COVID-19 Science Program Working Groups and Associated Chairs

- COVID-19 Related PheWAS
  - Katherine Liao, M.D.
    VA Boston Healthcare System, 150 S. Huntington Avenue, Boston, MA 02130

    - Scott Damrauer, M.D.
      Philadelphia VA Medical Center, 3900 Woodland Avenue, Philadelphia, PA 19104
- Disease Mechanisms
    - Richard Hauger, M.D.
      VA San Diego Healthcare System, 3350 La Jolla Village Drive, San Diego, CA 92161
    - Shiuh-Wen Luoh, M.D., Ph.D.
      Portland VA Medical Center, 3710 SW U.S. Veterans Hospital Road, Portland, OR 97239
    - Sudha Iyengar, Ph.D.
      VA Northeast Ohio Healthcare System, 10701 East Boulevard, Cleveland, OH 44106
- Druggable Genome
    - Juan P. Casas, M.D., Ph.D.
      VA Boston Healthcare System, 150 S. Huntington Avenue, Boston, MA 02130
- Genomics for Risk Prediction, PRS, and MR
    - Themistocles Assimes, M.D., Ph.D.
      VA Palo Alto Health Care System, 3801 Miranda Avenue, Palo Alto, CA 94304
    - Panagiotis Roussos, M.D., Ph.D.
      James J. Peters VA Medical Center, 130 W Kingsbridge Rd, Bronx, NY 10468
    - Robert Striker, M.D., Ph.D.
      William S. Middleton Memorial Veterans Hospital, 2500 Overlook Terrace, Madison, WI 53705
- GWAS & Downstream Analysis
    - Jennifer Huffman, Ph.D.
      VA Boston Healthcare System, 150 S. Huntington Avenue, Boston, MA 02130
    - Yan Sun, Ph.D.
      Atlanta VA Medical Center, 1670 Clairmont Road, Decatur, GA 30033
- Pharmacogenomics
    - Adriana Hung, M.D., M.P.H.
      VA Tennessee Valley Healthcare System, 1310 24th Avenue, South Nashville, TN 37212
    - Sony Tuteja, Pharm.D., M.S.
      Philadelphia VA Medical Center, 3900 Woodland Avenue, Philadelphia, PA 19104
- VA COVID-19 Shared Data Resource – Scott L. DuVall, Ph.D.; Kristine E. Lynch, Ph.D.; Elise Gatsby, M.P.H.
  VA Informatics and Computing Infrastructure (VINCI), VA Salt Lake City Health Care System, 500 Foothill Drive, Salt Lake City, UT 84148
- MVP COVID-19 Data Core – Kelly Cho, M.P.H., Ph.D.; Lauren Costa, M.P.H.; Anne Yuk-Lam Ho, M.P.H.; Rebecca Song, M.P.H.
  VA Boston Healthcare System, 150 S. Huntington Avenue, Boston, MA 02130

#### MVP COVID-19 Genomics and PRS Working Group Roster

- Panagiotis Roussos (Co-chair)
- Tim Assimes (Co-chair)
- Rob Striker (Co-chair)
- Helene Garcon (Initial co-ordinator)
- Jenny Huffman
- Jacob Joseph
- Reid Thompson
- Wen-Chih Wu
- Austin Nguyen
- Emily Wan
- Renato Polimanti
- Frank Wendt
- Dan Levey
- Pradeep Natarajan
- John McGeary
- Merry-Lynn McDonald
- Rob Igo
- Murray B Stein
- Michael Lewis
- Scott Damrauer
- Anurag Verma
- Kyong-Mi Chang
- Ravi Madduri
- Jimmy Efird
- Darshana Jhala
- Jeffrey Petersen
- James Meigs
- Benjamin Voight
- Joel Gelernter
- Jin Zhou
- Michael Levin
- Edward Siew
- Giorgio Sirugo
- Austin Hilliard
- Elvis Akwo
- Adriana Hung
- Shiuh-Wen Luoh
- Jessica Minnier
- Rose Huang
- Liam Gaziano

- Sharvari Dalal
- Cassy Robinson-Cohen
- Ran Tao
- Phil Tsao
- Mehrdad Arjomandi
- Marijana Vujkovic
- Anoop Sendamarai
- Uma Saxena
- Saiju Pyarajan

#### MVP COVID-19 GWAS Working Group Roster

- Jenny Huffman (Co-chair)
- Yan Sun (Co-chair)
- Helene Garcon (Initial co-ordinator)
- Jacob Joseph
- Reid Thompson
- Mary Wood
- Dana Crawford
- Emily Wan
- Renato Polimanti
- Frank Wendt
- Gita Pathak
- Pradeep Natarajan
- John McGeary
- Merry-Lynn McDonald
- Rob Igo
- Scott Damrauer
- Anurag Verma
- Ravi Madduri
- Jimmy Efird
- Benjamin Voight
- Joel Gelernter
- Michael Levin
- Richard Hauger
- Valerio Napolioni
- Michael Matheny
- Alex Bick
- Edward Siew
- Austin Hilliard
- Adriana Hung
- Shiuh-Wen Luoh
- Rose Huang

- Matt Freiberg
- Quinn S. Wells
- Cassy Robinson-Cohen
- Alex Pereira
- Kyong-Mi Chang
- Saiju Pyarajan
- Anoop Sendamarai
- Uma Saxena
- Gina Peloso
- Catherine Tcheandjieu

#### MVP Executive Committee

- Co-Chair: J. Michael Gaziano, M.D., M.P.H.
  VA Boston Healthcare System, 150 S. Huntington Avenue, Boston, MA 02130
- Co-Chair: Sumitra Muralidhar, Ph.D.
  US Department of Veterans Affairs, 810 Vermont Avenue NW, Washington, DC 20420
- Rachel Ramoni, D.M.D., Sc.D., Chief VA Research and Development Officer
  US Department of Veterans Affairs, 810 Vermont Avenue NW, Washington, DC 20420
- Jean Beckham, Ph.D.
  Durham VA Medical Center, 508 Fulton Street, Durham, NC 27705
- Kyong-Mi Chang, M.D.
  Philadelphia VA Medical Center, 3900 Woodland Avenue, Philadelphia, PA 19104
- Christopher J. O'Donnell, M.D., M.P.H.
  VA Boston Healthcare System, 150 S. Huntington Avenue, Boston, MA 02130
- Philip S. Tsao, Ph.D.
  VA Palo Alto Health Care System, 3801 Miranda Avenue, Palo Alto, CA 94304
- James Breeling, M.D., Ex-Officio
  US Department of Veterans Affairs, 810 Vermont Avenue NW, Washington, DC 20420
- Grant Huang, Ph.D., Ex-Officio
  US Department of Veterans Affairs, 810 Vermont Avenue NW, Washington, DC 20420
- Juan P. Casas, M.D., Ph.D., Ex-Officio
  VA Boston Healthcare System, 150 S. Huntington Avenue, Boston, MA 02130

#### MVP Program Office

- Sumitra Muralidhar, Ph.D.
  US Department of Veterans Affairs, 810 Vermont Avenue NW, Washington, DC 20420
- Jennifer Moser, Ph.D.
  US Department of Veterans Affairs, 810 Vermont Avenue NW, Washington, DC 20420

#### MVP Recruitment/Enrollment

- Recruitment/Enrollment Director/Deputy Director, Boston – Stacey B. Whitbourne, Ph.D.; Jessica V. Brewer, M.P.H.
  VA Boston Healthcare System, 150 S. Huntington Avenue, Boston, MA 02130
- MVP Coordinating Centers
  - Clinical Epidemiology Research Center (CERC), West Haven – Mihaela Aslan, Ph.D.
    West Haven VA Medical Center, 950 Campbell Avenue, West Haven, CT 06516
  - Cooperative Studies Program Clinical Research Pharmacy Coordinating Center, Albuquerque – Todd Connor, Pharm.D.; Dean P. Argyres, B.S., M.S.
    New Mexico VA Health Care System, 1501 San Pedro Drive SE, Albuquerque, NM 87108
  - Genomics Coordinating Center, Palo Alto – Philip S. Tsao, Ph.D.
    VA Palo Alto Health Care System, 3801 Miranda Avenue, Palo Alto, CA 94304
  - MVP Boston Coordinating Center, Boston - J. Michael Gaziano, M.D., M.P.H.
    VA Boston Healthcare System, 150 S. Huntington Avenue, Boston, MA 02130
  - MVP Information Center, Canandaigua – Brady Stephens, M.S.
    Canandaigua VA Medical Center, 400 Fort Hill Avenue, Canandaigua, NY 14424
- VA Central Biorepository, Boston – Mary T. Brophy M.D., M.P.H.; Donald E. Humphries, Ph.D.; Luis E. Selva, Ph.D.
  VA Boston Healthcare System, 150 S. Huntington Avenue, Boston, MA 02130
- MVP Informatics, Boston – Nhan Do, M.D.; Shahpoor (Alex) Shayan, M.S.
  VA Boston Healthcare System, 150 S. Huntington Avenue, Boston, MA 02130
- MVP Data Operations/Analytics, Boston – Kelly Cho, M.P.H., Ph.D.
  VA Boston Healthcare System, 150 S. Huntington Avenue, Boston, MA 02130
- Director of Regulatory Affairs – Lori Churby, B.S.
  VA Palo Alto Health Care System, 3801 Miranda Avenue, Palo Alto, CA 94304

#### MVP Science

- Science Operations – Christopher J. O'Donnell, M.D., M.P.H.
  VA Boston Healthcare System, 150 S. Huntington Avenue, Boston, MA 02130
- Genomics Core
  - Christopher J. O'Donnell, M.D., M.P.H.
    VA Boston Healthcare System, 150 S. Huntington Avenue, Boston, MA 02130
  - Saiju Pyarajan Ph.D.
    VA Boston Healthcare System, 150 S. Huntington Avenue, Boston, MA 02130
  - Philip S. Tsao, Ph.D.
    VA Palo Alto Health Care System, 3801 Miranda Avenue, Palo Alto, CA 94304
- Data Core - Kelly Cho, M.P.H, Ph.D.
  VA Boston Healthcare System, 150 S. Huntington Avenue, Boston, MA 02130

- VA Informatics and Computing Infrastructure (VINCI) – Scott L. DuVall, Ph.D.
  VA Salt Lake City Health Care System, 500 Foothill Drive, Salt Lake City, UT 84148
- Data and Computational Sciences – Saiju Pyarajan, Ph.D.
  VA Boston Healthcare System, 150 S. Huntington Avenue, Boston, MA 02130
- Statistical Genetics
  - Elizabeth Hauser, Ph.D.
    Durham VA Medical Center, 508 Fulton Street, Durham, NC 27705
  - Yan Sun, Ph.D.
    Atlanta VA Medical Center, 1670 Clairmont Road, Decatur, GA 30033
  - Hongyu Zhao, Ph.D.
    West Haven VA Medical Center, 950 Campbell Avenue, West Haven, CT 06516

#### Current MVP Local Site Investigators

- Atlanta VA Medical Center (Peter Wilson, M.D.)
  1670 Clairmont Road, Decatur, GA 30033
- Bay Pines VA Healthcare System (Rachel McArdle, Ph.D.)
  10,000 Bay Pines Blvd Bay Pines, FL 33744
- Birmingham VA Medical Center (Louis Dellitalia, M.D.)
  700 S. 19th Street, Birmingham AL 35233
- Central Western Massachusetts Healthcare System (Kristin Mattocks, Ph.D., M.P.H.)
  421 North Main Street, Leeds, MA 01053
- Cincinnati VA Medical Center (John Harley, M.D., Ph.D.)
  3200 Vine Street, Cincinnati, OH 45220
- Clement J. Zablocki VA Medical Center (Jeffrey Whittle, M.D., M.P.H.)
  5000 West National Avenue, Milwaukee, WI 53295
- VA Northeast Ohio Healthcare System (Frank Jacono, M.D.)
  10701 East Boulevard, Cleveland, OH 44106
- Durham VA Medical Center (Jean Beckham, Ph.D.)
  508 Fulton Street, Durham, NC 27705
- Edith Nourse Rogers Memorial Veterans Hospital (John Wells., Ph.D.)
  200 Springs Road, Bedford, MA 01730
- Edward Hines, Jr. VA Medical Center (Salvador Gutierrez, M.D.)
  5000 South 5th Avenue, Hines, IL 60141
- Veterans Health Care System of the Ozarks (Gretchen Gibson, D.D.S., M.P.H.)
  1100 North College Avenue, Fayetteville, AR 72703
- Fargo VA Health Care System (Kimberly Hammer, Ph.D.)
  2101 N. Elm, Fargo, ND 58102
- VA Health Care Upstate New York (Laurence Kaminsky, Ph.D.)
  113 Holland Avenue, Albany, NY 12208
- New Mexico VA Health Care System (Gerardo Villareal, M.D.)
  1501 San Pedro Drive, S.E. Albuquerque, NM 87108

- VA Boston Healthcare System (Scott Kinlay, M.B.B.S., Ph.D.)
  150 S. Huntington Avenue, Boston, MA 02130
- VA Western New York Healthcare System (Junzhe Xu, M.D.)
  3495 Bailey Avenue, Buffalo, NY 14215-1199
- Ralph H. Johnson VA Medical Center (Mark Hamner, M.D.)
  109 Bee Street, Mental Health Research, Charleston, SC 29401
- Columbia VA Health Care System (Roy Mathew, M.D.)
  6439 Garners Ferry Road, Columbia, SC 29209
- VA North Texas Health Care System (Sujata Bhushan, M.D.)
  4500 S. Lancaster Road, Dallas, TX 75216
- Hampton VA Medical Center (Pran Iruvanti, D.O., Ph.D.)
  100 Emancipation Drive, Hampton, VA 23667
- Richmond VA Medical Center (Michael Godschalk, M.D.)
  1201 Broad Rock Blvd., Richmond, VA 23249
- Iowa City VA Health Care System (Zuhair Ballas, M.D.)
  601 Highway 6 West, Iowa City, IA 52246-2208
- Eastern Oklahoma VA Health Care System (Douglas Ivins, M.D.)
  1011 Honor Heights Drive, Muskogee, OK 74401
- James A. Haley Veterans' Hospital (Stephen Mastorides, M.D.)
  13000 Bruce B. Downs Blvd, Tampa, FL 33612
- James H. Quillen VA Medical Center (Jonathan Moorman, M.D., Ph.D.)
  Corner of Lamont & Veterans Way, Mountain Home, TN 37684
- John D. Dingell VA Medical Center (Saib Gappy, M.D.)
  4646 John R Street, Detroit, MI 48201
- Louisville VA Medical Center (Jon Klein, M.D., Ph.D.)
  800 Zorn Avenue, Louisville, KY 40206
- Manchester VA Medical Center (Nora Ratcliffe, M.D.)
  718 Smyth Road, Manchester, NH 03104
- Miami VA Health Care System (Hermes Florez, M.D., Ph.D.)
  1201 NW 16th Street, 11 GRC, Miami FL 33125
- Michael E. DeBakey VA Medical Center (Olaoluwa Okusaga, M.D.)
  2002 Holcombe Blvd, Houston, TX 77030
- Minneapolis VA Health Care System (Maureen Murdoch, M.D., M.P.H.)
  One Veterans Drive, Minneapolis, MN 55417
- N. FL/S. GA Veterans Health System (Peruvemba Sriram, M.D.)
  1601 SW Archer Road, Gainesville, FL 32608
- Northport VA Medical Center (Shing Shing Yeh, Ph.D., M.D.)
  79 Middleville Road, Northport, NY 11768
- Overton Brooks VA Medical Center (Neeraj Tandon, M.D.)
  510 East Stoner Ave, Shreveport, LA 71101
- Philadelphia VA Medical Center (Darshana Jhala, M.D.)
  3900 Woodland Avenue, Philadelphia, PA 19104

- Phoenix VA Health Care System (Samuel Aguayo, M.D.)
  650 E. Indian School Road, Phoenix, AZ 85012
- Portland VA Medical Center (David Cohen, M.D.)
  3710 SW U.S. Veterans Hospital Road, Portland, OR 97239
- Providence VA Medical Center (Satish Sharma, M.D.)
  830 Chalkstone Avenue, Providence, RI 02908
- Richard Roudebush VA Medical Center (Suthat Liangpunsakul, M.D., M.P.H.)
  1481 West 10th Street, Indianapolis, IN 46202
- Salem VA Medical Center (Kris Ann Oursler, M.D.)
  1970 Roanoke Blvd, Salem, VA 24153
- San Francisco VA Health Care System (Mary Whooley, M.D.)
  4150 Clement Street, San Francisco, CA 94121
- South Texas Veterans Health Care System (Sunil Ahuja, M.D.)
  7400 Merton Minter Boulevard, San Antonio, TX 78229
- Southeast Louisiana Veterans Health Care System (Joseph Constans, Ph.D.)
  2400 Canal Street, New Orleans, LA 70119
- Southern Arizona VA Health Care System (Paul Meyer, M.D., Ph.D.)
  3601 S 6th Avenue, Tucson, AZ 85723
- Sioux Falls VA Health Care System (Jennifer Greco, M.D.)
  2501 W 22nd Street, Sioux Falls, SD 57105
- St. Louis VA Health Care System (Michael Rauchman, M.D.)
  915 North Grand Blvd, St. Louis, MO 63106
- Syracuse VA Medical Center (Richard Servatius, Ph.D.)
  800 Irving Avenue, Syracuse, NY 13210
- VA Eastern Kansas Health Care System (Melinda Gaddy, Ph.D.)
  4101 S 4th Street Trafficway, Leavenworth, KS 66048
- VA Greater Los Angeles Health Care System (Agnes Wallbom, M.D., M.S.)
  11301 Wilshire Blvd, Los Angeles, CA 90073
- VA Long Beach Healthcare System (Timothy Morgan, M.D.)
  5901 East 7th Street Long Beach, CA 90822
- VA Maine Healthcare System (Todd Stapley, D.O.)
  1 VA Center, Augusta, ME 04330
- VA New York Harbor Healthcare System (Scott Sherman, M.D., M.P.H.)
  423 East 23rd Street, New York, NY 10010
- VA Pacific Islands Health Care System (George Ross, M.D.)
  459 Patterson Rd, Honolulu, HI 96819
- VA Palo Alto Health Care System (Philip Tsao, Ph.D.)
  3801 Miranda Avenue, Palo Alto, CA 94304-1290
- VA Pittsburgh Health Care System (Patrick Strollo, Jr., M.D.)
  University Drive, Pittsburgh, PA 15240
- VA Puget Sound Health Care System (Edward Boyko, M.D.)
  1660 S. Columbian Way, Seattle, WA 98108-1597

- VA Salt Lake City Health Care System (Laurence Meyer, M.D., Ph.D.)
  500 Foothill Drive, Salt Lake City, UT 84148
- VA San Diego Healthcare System (Samir Gupta, M.D., M.S.C.S.)
  3350 La Jolla Village Drive, San Diego, CA 92161
- VA Sierra Nevada Health Care System (Mostaqul Huq, Pharm.D., Ph.D.)
  975 Kirman Avenue, Reno, NV 89502
- VA Southern Nevada Healthcare System (Joseph Fayad, M.D.)
  6900 North Pecos Road, North Las Vegas, NV 89086
- VA Tennessee Valley Healthcare System (Adriana Hung, M.D., M.P.H.)
  1310 24th Avenue, South Nashville, TN 37212
- Washington DC VA Medical Center (Jack Lichy, M.D., Ph.D.)
  50 Irving St, Washington, D. C. 20422
- W.G. (Bill) Hefner VA Medical Center (Robin Hurley, M.D.)
  1601 Brenner Ave, Salisbury, NC 28144
- White River Junction VA Medical Center (Brooks Robey, M.D.)
  163 Veterans Drive, White River Junction, VT 05009
- William S. Middleton Memorial Veterans Hospital (Robert Striker, M.D., Ph.D.)
  2500 Overlook Terrace, Madison, WI 53705

### Mount Sinai COVID-19 Biobank: Core Acknowledgment for Publications

Icahn School of Medicine at Mount Sinai, New York, NY, 10029, USA

Charuta Agashe, Priyal Agrawal, Alara Akyatan, Kasey Alesso-Carra, Eziwoma Alibo, Kelvin Alvarez, Angelo Amabile, Carmen Argmann, Kimberly Argueta, Steven Ascolillo, Rasheed Bailey, Craig Batchelor, Noam D. Beckmann, Aviva G. Beckmann, Priya Begani, Jessica Le Berichel, Dusan Bogunovic, Swaroop Bose, Cansu Cimen Bozkus, Paloma Bravo, Mark Buckup, Larissa Burka, Sharlene Calorossi, Lena Cambron, Guillermo Carbonell, Gina Carrara, Mario A. Cedillo, Christie Chang, Serena Chang, Alexander W. Charney, Steven T. Chen, Esther Cheng, Jonathan Chien, Mashkura Chowdhury, Jonathan Chung, Phillip H. Comella, Dana Cosgrove, Francesca Cossarini, Liam Cotter, Arpit Dave, Travis Dawson, Bheesham Dayal, Diane Marie Del Valle, Maxime Dhainaut, Rebecca Dornfeld, Katie Dul, Melody Eaton, Nissan Eber, Cordelia Elaiho, Ethan Ellis, Frank Fabris, Jeremiah Faith, Dominique Falci, Susie Feng, Brian Fennessy, Marie Fernandes, Nataly Fishman, Nancy J. Francoeur, Sandeep Gangadharan, Daniel Geanon, Bruce D. Gelb, Benjamin S. Glicksberg, Sacha Gnjatic, Joanna Grabowska, Gavin Gyimesi, Maha Hamdani, Diana Handler, Jocelyn Harris, Matthew Hartnett, Sandra Hatem, Manon Herbinet, Elva Herrera, Arielle Hochman, Gabriel E. Hoffman, Jaime Hook, Laila Horta, Etienne Humblin, Suraj Jaladanki, Hajra Jamal, Jessica S. Johnson, Gurpawan Kang, Neha Karekar, Subha Karim, Geoffrey Kelly, Jong Kim, Seunghee Kim-Schulze, Arvind Kumar, Jose Lacunza, Alona Lansky, Dannielle Lebovitch, Brian Lee, Grace Lee, Gyu Ho Lee, Jacky Lee, John Leech, Lauren Lepow, Michael B. Leventhal, Lora E. Liharska, Katherine Lindblad, Alexandra Livanos, Bojan Losic, Rosalie Machado, Kent Madrid, Zafar Mahmood, Kelcey Mar, Thomas U. Marron, Glenn Martin, Robert Marvin, Shrisha Maskey, Paul Matthews, Katherine Meckel, Saurabh Mehandru, Miriam Merad, Cynthia Mercedes, Elyze Merzier, Dara Meyer, Gurkan Mollaoglu, Sarah Morris, Konstantinos Mouskas, Emily Moya, Naa-akomaah Yeboah, Girish Nadkarni, Kai Nie, Marjorie Nisenholtz, George Ofori-Amanfo, Kenan Onel, Merouane Ounadjela, Manishkumar Patel, Vishwendra Patel, Cassandra Pruitt, Adeeb Rahman, Shivani Rathi, Jamie Redes, Ivan Reyes-Torres, Alcina Rodrigues, Alfonso Rodriguez, Vladimir Roudko, Panos Roussos, Evelyn Ruiz, Pearl Scalzo, Eric E. Schadt, Ieisha Scott, Robert Sebra, Hardik Shah, Pedro Silva, Nicole W. Simons, Melissa Smith, Alessandra Soares Schanoski, Juan Soto, Shwetha Hara Sridhar, Stacey-Ann Brown, Hiyab Stefanos, Meghan Straw, Robert Sweeney, Alexandra Tabachnikova, Collin Teague, Ryan Thompson, Manying Tin, Kevin Tuballes, Scott R. Tyler, Bhaskar Upadhyaya, Akhil Vaid, Verena Van Der Heide, Natalie Vaninov, Konstantinos Vlachos, Daniel Wacker, Laura Walker, Hadley Walsh, Bo Wang, Wenhui Wang, C. Matthias Wilk, Lillian Wilkins, Karen M. Wilson, Jessica Wilson, Hui Xie, Li Xue, Nancy Yi, Ying-chih Wang, Mahlet Yishak, Sabina Young, Alex Yu, Nina Zaks and Renyuan Zha.

#### Other Acknowledgements

Figure 4B: Created with BioRender.com.

### SUPPLEMENTARY MATERIALS AND METHODS

### Transcriptome-wide association study

GWAS summary statistics were obtained from https://www.covid19hg.org/results/r4/; we use the phenotypes and files detailed in Supplementary Table 2 from the 4th release (2020-10-20).

### Genetically regulated gene expression (GReX-) based gene targeting approach and computational drug repurposing

**Perturbargen Library used.** We use the LINKS Phase II L1000 dataset (GSE70138) perturbagen reference library[1]. All inferred genes (AIG; n=12,328) are considered. Only "gold" signatures are considered. **Imputed transcriptomes used.** Analysis is limited to 17 imputed transcriptomes: (1) the B2 phenotype, and (2) EpiXcan tissue models that have at least one FDR-significant finding (FDR significance takes into account all COVID phenotypes and all 42 tissue models considered): Adipose: subcutaneous (GTEx), Adipose: subcutaneous (STARNET), Adipose: visceral (GTEx), Adipose: visceral (STARNET), Artery: Aorta (GTEx), Artery: Aorta (STARNET), Artery: Mammary (STARNET), Blood (STARNET), GI: esophagus, GE junction (GTEx), GI: esophagus, mucosa (GTEx), GI: muscularis (GTEx), GI: pancreas (GTEx), Muscle: skeletal (GTEx), Muscle: skeletal (STARNET), Reproductive: mammary tissue (GTEx), Respiratory: lung (GTEx), Skin: sun exposed lower leg (GTEx). **Summarization and prioritization approach:** From the original CDR pipeline[2] (using 100 permutations), applied to the imputed transcriptome of each tissue, we obtain the average rank of the compound in antagonizing the GREx and the permutation p values. After pulling together all the results, we perform a Mann-Whithney U test for each candidate compound/ shRNA against all other compounds to see if the candidate's rankings significantly deviate from the median rank. For each candidate we also estimate a GREx antagonism pseudo z-score, which is defined as the negative Hodges-Lehmann estimator (of the median difference between that specific candidate vs. the other candidates) divided by the standard deviation of the ranks of the compounds ($-\frac{Hodges-Lehmann\ estimator_{perturbagen}}{SD\ average\ ranks\ of\ all\ perturbagens}$); a positive pseudo z-score is interpreted as a potential therapeutic candidate whereas a negative pseudo z-score would suggest that the shRNA is not antagonizing the imputed transcriptome and is thus likely to exacerbate the phenotype. Of note is that at this stage each candidate is compared against the other candidates but we can confirm that the candidate is effectively antagonizing the GREx by looking at the original permutation p values. FDR is estimated with the Benjamini–Hochberg procedure[3]. **Additional information for chemical compound analyses:** Analysis is limited to compounds eligible for drug repurposing (n=495). Drug information for the compounds under consideration (e.g. clinical phase, mechanism of action and molecular targets) was obtained from http://www.broadinstitute.org/repurposing (file date: 3/24/2020). For comparison with other studies; the compounds under question were compared with all the other compounds. For the mechanism of action comparison all compounds with a known mechanism of action represented with two or more candidates are considered. Final recommendations are for launched medications and FDR correction is applied only to launched compounds. **Additional information for shRNA analyses:** All shRNAs were considered.

#### Gene expression profiling and EHR-based phenotyping in the Mount Sinai COVID-19 Biobank (full methods, dataset and results will be presented in Beckmann et al, manuscript in preparation).

RNA extraction, RNA-seq library preparation and sequencing, and RNA-seq data processing, quality control and normalization as previously described[4]. In addition, we confirmed that no samples were mislabeled. After removing lowly expressed genes (keeping genes with counts per million > 1 in 10% of the samples), we normalized the raw count data of the 21,114 remaining genes using voom from the limma R package[5]. Samples that failed to pass all quality controls were removed from further analyses. Principal components analyses to explore covariate effect on gene expression variance genome-wide were done using the prcomp R function. Batch effect was calculated on a per gene basis using technical replicates sequenced in all batches. The following additional covariates were included in the model: Subject ID, number of days since first blood sample, RNA Library Prep Plate, Sex, DV200 Percent and PICARD metrics PCT_R2_TRANSCRIPT_STRAND_READS, PCT_INTRONIC_BASES, WIDTH_OF_95_PERCENT, MEDIAN_5PRIME_BIAS, MEDIAN_3PRIME_BIAS. **Cell-type deconvolution.** Cell type deconvolution was performed with the Cibersortx software, using transcripts per million as input and following procedures recommended by the developers[6] using 3 independently generated references[7,8] as previously described[9]. Cell types relevant to severity were defined with a linear mixed model lasso procedure (R package glmmlasso). **Differential Expression.** Differential expression (DE) analyses were performed with the dream function from the variancePartition R package[10,11]. The covariates included in the model are described aboveA total of 6 DE signatures were generated (Severe end-organ damage Vs. Severe; Severe end-organ damage Vs. Moderate; Severe end-organ damage Vs. Control; Severe Vs. Moderate; Severe Vs. Control; Moderate Vs. Control). Multiple testing was controlled separately for each DE comparison accounting for the 21,114 genes tested using the false discovery rate (FDR) estimation method of Benjamini-Hochberg[3].

#### Manipulation of IL10RB and IFNAR2 expression in cell lines and their effect on SARS-CoV-2 viral load and transcriptional profiles

**hiPSC-NPC culture and donor.** hiPSC-NPCs of line NSB2607 (male, 15 years old, European descent)[12] were cultured in hNPC media (DMEM/F12 (Life Technologies #10565), 1x N-2 (Life Technologies #17502-048), 1x B-27-RA (Life Technologies #12587-010), 20 ng/mL FGF2 (Life Technologies)) on Matrigel (Corning, #354230). hiPSC-NPCs at full confluence (1-1.5x$10^7$ cells / well of a 6-well plate) were dissociated with Accutase (Innovative Cell Technologies) for 5 mins, spun down (5 mins X 1000g), resuspended and seeded onto Matrigel-coated plates at 3-5x$10^6$ cells / well. Media was replaced every 24 days for 4 to 7 days until the next split.

**SARS-CoV-2 virus propagation and infections.** SARS-related coronavirus 2 (SARS-CoV-2), isolate USA-WA1/2020 (NR-52281) was deposited by the Center for Disease Control and Prevention and obtained through BEI Resources, NIAID, NIH. SARS-CoV-2 was propagated in Vero E6 cells in DMEM supplemented with 2% FBS, 4.5 g/L D-glucose, 4 mM L-glutamine, 10 mM Non-Essential Amino Acids, 1 mM Sodium Pyruvate and 10 mM HEPES. Virus stock was filtered by centrifugation using Amicon Ultra-15 Centrifugal filter unit (Sigma, Cat # UFC910096) and resuspended in viral propagation media. All infections were performed with either passage 3 or 4 SARS-CoV-2. Infectious titers of SARS-CoV-2 were determined by plaque assay in Vero E6 cells in Minimum Essential Media supplemented with 4mM L-glutamine, 0.2% BSA, 10 mM HEPES and 0.12% NaHCO3 and 0.7% Oxoid agar (Cat #OXLP0028B). All SARS-CoV-2 infections were performed in the CDC/USDA-approved BSL-3 facility of the Global Health and Emerging Pathogens Institute at the Icahn School of Medicine at Mount Sinai in accordance with institutional biosafety requirements.

**gRNA Design and Cloning and shRNAs.** gRNAs were designed using the CRISPR-ERA (http://crispr-era.stanford. edu) web tool. gRNAs were selected based on their specific locations at decreasing distances from the TSS as well as their lack of predicted off targets and E scores (http://crispr-era.stanford.edu). For lentiviral cloning: synthesized oligonucleotides were phospho-annealed (37°C for 30 min, 95°C for 5 min, ramped-down to 25°C at 5°C per min), diluted 1:100 and then ligated into BsmB1-digested lentiGuide-Hygro-mTagBFP2 (addgene Plasmid #99374). Product was transformed into NEB10-beta E. coli. via manufacturer's protocol (NEB # C3019H). shRNAs were ordered as glycerol stocks from Sigma (IL10RB # SHCLNG-NM_000628; IFNAR2 # SHCLNG-NM_000874). **Gibson Assembly of Vectors:** Unless specified, all cloning reagents were from NEB and plasmid backbones were from Addgene (https://www.addgene.org/). Primers were synthesized by Thermo Fisher Scientific. All fragments were assembled using NEBuilder HiFi DNA Assembly Master Mix (NEB, no. E2621X). All assemblies were transformed into either DH5a Extreme Efficiency Competent Cells (Allele Biotechnology, no. ABP-CE-CC02050) or Stbl3 Chemically Competent E. coli (Thermo Fisher Scientific, no. C737303). Positive clones were confirmed by restriction digest and Sanger sequencing (GENEWIZ). The following vectors have been deposited at Addgene: lenti-EF1a- dCas9-VP64-Puro, lenti-EF1a-dCas9-VPR-Puro, lenti-EF1a-dCas9-KRAB-Puro, lentiGuide-Hygro-mTagBFP2, lentiGuide-Hygro-eGFP, lentiGuide-Hygro-dTomato, lentiGuide-Hygro-iRFP670, and pLV-TetO-hNGN2-Neo. lentiGuide-dTomato and lentiGuide-mTagBFP2-Hygro lentiGuide-Puro (Addgene, no. 52963) were digested with Mlu1 and BsiWI. dTomato was amplified from AAV-hSyn1-GCaMP6f-P2A-NLS-dTomato (Addgene, no.51085). HygroR sequence was amplified from lentiMS2-P65-HSF1_Hygro (Addgene, no. 61426). mTagBFP2 was amplified from pBAD-mTagBFP2 (Addgene, no. 3463). The P2A self-cleaving peptide sequence was amplified using a reverse primer of HygroR and forward primer of mTagBFP2. All gRNA sequences are provided in Supplementary Table 16.

**Lentiviral dCas9 Effectors.** To engineer a lentiviral transfer vector that expresses dCas9: VP64-T2A-Puro (EF1a-NLS-dCas9(N863)-VP64-T2A-Puro-WPRE), dCas9:VP64-T2A-Blast (EF1a-NLS-dCas9(N863)-VP64-T2A-Blast-WPRE) (Addgene, no. 61,425) was digested with BsrGI and EcoRI. T2A-PuroR was amplified from pLV-TetO-hNGN2-P2A-eGFP-T2A-Puro (Addgene, no. 79823). Fragments were then assembled using NEBuilder HiFi DNA Assembly

Master Mix (NEB, no. E2621). To engineer a lentiviral transfer vector that expresses dCas9:KRAB-Puro (EF1a-NLS-dCas9(N863)-KRAB-T2A-Puro-WPRE), dCas9:VP64-T2A-Blast (EF1a-NLS-dCas9(N863)-VP64-T2A-Blast-WPRE) (Addgene, no.61425) was first digested with BamHI and BsrGI. KRAB was then amplified from pHAGE-TRE-dCas9:KRAB (Addgene, no. 50917). Fragments were then assembled using NEBuilder HiFi DNA Assembly Master Mix. dCas9:KRAB-Blast was digested with BsrGI and EcoRI, and T2A-PuroR was amplified from pLV-TetO-hNGN2- P2A-eGFP-T2A-Puro (Addgene, no. 79823). Fragments were then assembled using NEBuilder HiFi DNA Assembly Master Mix. To engineer a lentiviral transfer vector that expresses dCas9:VPR-Puro (EF1a- NLS-dCas9(D10A, D839A, H840A, and N863A)-VPR-T2A-Puro- WPRE), dCas9:VPR was first amplified from SP-dCas9-VPR (Addgene, no. 63798), and T2A-PuroR was amplified from pLV-TetO-hNGN2-P2A-eGFP-T2A-Puro (Addgene, no. 79823). dCas9:KRAB-T2A-Puro was digested with BsiWI and EcoRI. Fragments were then assembled using NEBuilder HiFi DNA Assembly Master Mix.

**NGN2-glutamatergic neuron induction of shRNA and CRISPRa treated neurons[13,14].** On day -1 NPCs were dissociated with Accutase Cell Detachment Solution for 5min at 37°C, counted and seeded at a density of at $5x10^5$ cells/well on Matrigel coated 24-well plates in hNPC media (DMEM/F12 (Life Technologies #10565), 1x N-2 (Life Technologies #17502-048), 1x B-27-RA, 20 ng/mL FGF2 (Life Technologies)) on Matrigel (Corning, #354230). On day 0, cells were transduced with rtTA and NGN2 lentiviruses as well as desired shRNA or CRISPRa viruses in NPC media containing 10 μM Thiazovivin and spinfected (centrifuged for 1 hour at 1000g). On day 1, media was replaced and doxycycline was added with 1ug/mL working concentration. On day 2, transduced hNPCs were treated with corresponding antibiotics to the lentiviruses (1 μg/mL puromycin for shRNA, 1 mg/mL G-418 for NGN2-Neo). On day 4, medium was switched to Brainphys neuron medium plus 1 μg/mL dox. Medium was replaced every second day until SARS-CoV-2 (MOI of 0.1) or mock infection on day 7. The samples were harvested in Trizol (Invitrogen, Cat #15596026) 24 hours later. RNA was isolated by phenol/chloroform extraction prior to purification using the RNeasy Mini Kit (Qiagen, Cat # 74106).

**Sequencing Platform.** RNA samples were received at the New York Genome Center and, followingan initial quality check, were normalized onto two different 96 well plates for a total RNA with RiboErase assay and a SARS-CoV-2 targeted assay. For the total RNA assay 200ng of RNA were normalized into a plate to be run through the KAPA RNA Hyper Prep Kit + RiboErase HMR (Roche, cat no: 8098140702). This total RNA prep followed the manufacturer's protocol with minor adjustments for automation on the PerkinElmer sciclone. Briefly, the RNA first goes through an oligo hybridization and rRNA depletion and then 1st and 2nd strand synthesis. The cDNA then gets adenylated ends and unique dual indexed adapters ligated onto the ends. Finally, the sample goes through a clean up followed by enrichment and purification. The final library was quantified by picogreen and ran on a fragment analyzer to determine final library size. Samples were normalized, pooled and run on a NovaSeq 6000 S4 in a 2x150 run format targeting 60M reads per sample. For the SARS-CoV-2 targeted assay we used the AmpliSeq Library Plus and cDNA Synthesis for Illumina kits (Illumina, Cat no: 20019103 & 20022654). Briefly, 20ng of RNA were reverse transcribed, the cDNA targets were then amplified with the Illumina SARS-CoV-2 research panel (Illumina, 20020496). The amplicons were partially digested and AmpliSeq CD Indexes were ligated onto the amplicons. The library was then cleaned up and amplified. After amplification there was a final clean up and the libraries were quantified, pooled and run on a NovaSeq 6000 S4 in a 2x150 run format.

**SARS-CoV-2 quantification.** Short-read data were taxonomically classified using taxMaps[15]. As part of the taxMaps pipeline, reads were processed prior to mapping. Adapter sequences and low quality (Q < 20) bases were trimmed out and low complexity reads discarded. The remaining reads were then concurrently mapped against 1) the phiX174 reference genome (NC_001422.1); 2) the SARS-CoV-2 reference genome (NC_045512.2); and 3) a combined index encompassing the entire NCBI's nt database, RefSeq archaeal, bacterial, fungal, protozoan and viral genomes, as well as a selection of RefSeq model organism genomes, including the human GRCh38 reference[16], to produce the final classification. Given that some human sequences of ancestral origin (that constitute variation between individuals) are absent from the GRCh38 reference, a small percentage of human reads usually maps to other primate genomes and, consequently, is classified as such. To obtain more accurate estimates of the human content in these samples, all reads classified as "primate" were considered of human origin and reclassified accordingly. SARS-CoV-2 viral load was determined as the number of SARS-CoV-2 reads over the host (human) reads.

**Competitive gene set testing.** Competitive gene set testing using sets from Gene Ontology[17] was performed with camera[18]. First we performed differential expression analysis with limma[5] using the first two components of multidimensional scaling and RIN as covariates to identify the signature of SARS-CoV-2 infection in our cells while adjusting for other treatments. We then performed competitive gene set enrichment analysis for all gene ontology and betacoronavirus gene sets (n=18,553). For gene ontology datasets, we kept all significantly enriched gene sets (FDR<0.05) and kept those with a Jaccard index less than 0.2. For the betacoronavirus gene sets we kept all the gene sets and filtered based on a Jaccard index of 0.2. The combined SARS-CoV-2 gene set collection with the two datasets above (significant pruned gene ontology and all pruned betacoronavirus) was used for all the following competitive gene set testing except as otherwise indicated (n=296). Thus, in Figure 4D, enrichment analysis is run across the whole exploratory dataset (n=18,553) for SARS-CoV-2 infection (first row) whereas for all other conditions we are only exploring the combined SARS-CoV-2 gene set collection (n=296).

### SUPPLEMENTARY RESULTS

#### COVID-19 phenotypes genetically regulated gene expression (GREx) comparison.

As shown by correlation, hierarchical and principal component analysis (Supplementary Figure 2) of the COVID-19 phenotypes GREx, the phenotypes mainly cluster in 4 groups: *(1)* The severe vs. not severe COVID group (A1 and B1), *(2)* The severe COVID vs. population group (A2 and B2), *(3)* the any COVID vs. population or lab/self-reported negative group (C1 and C2) and finally *(4)* a group comprising the predicted COVID phenotype (D1). Of note is that this GREx-based phenotypic clustering persists despite differences in the different ancestries included in the genetic analysis (e.g. A1&B1, A2&B2, C1&C2) (Supplementary Table 2).

### SUPPLEMENTARY FIGURES

### Supplementary Figure 1

#### A. Signature vs. GREx (genetically regulated gene expression) antagonism (compounds/shRNA)

For each tissue TWAS (imputed GREx). Example: EpiXcan model of lung tissue (GTEx) for B2 COVID-19 phenotype

For each signature. Example: KDA010_MCF7_96H:TRCN0000058267:-666
Example is with shRNA but approach is identical for other compounds or perturbagens.

Cells: MCF7; shRNA: IL10RB; Tx duration: 96 hours

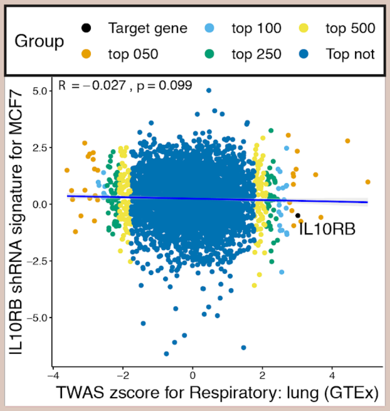


Five method rank approach (as per So et al. 2017) for each signature

For each signature and genetically regulated gene expression (GREx) from a single tissue:
- For top 50, 100, 250 and 500 differentially expressed TWAS genes perform:
  1. Pearson correlation
  2. Spearman correlation
- For all genes perform:
  3. Kolmogorov-Smirnov test (performed separately for top 50, 100, 250 and 500 DEG TWAS genes)
  4. Pearson correlation
  5. Spearman correlation

- Permutation tests (n=100) for each signature-TWAS pair by shuffling the TWAS z-scores and comparing them to signatures. For a compound to be considered therapeutic there must be at least one signature with p<=0.05 with the permutation method

Get average rank and other metrics from the five method rank approach (vs. all other signatures) for next step.

After per signature analysis is done for each tissue, all results from the five method rank approach are pulled together.

#### B. Summarization of effect of signatures at the level of compound/shrna for all cell lines and tissues

| Perturbagen | Cell line | TWAS tissue | Average Rank |
|---|---|---|---|
| IL10RB | Cell line 1 | Tissue 1 | 10 |
| GENE 1 | Cell line 2 | Tissue 1 | 15 |
| GENE 2 | Cell line 1 | Tissue 1 | 25 |
| IL10RB | Cell line 2 | Tissue 2 | 35 |
| IL10RB | Cell line 2 | Tissue 1 | 38 |
| GENE 2 | Cell line 2 | Tissue 1 | 40 |
| IL10RB | Cell line 1 | Tissue 2 | 42 |
| GENE 1 | Cell line 1 | Tissue 1 | 47 |
| GENE 1 | Cell line 2 | Tissue 2 | 50 |
| GENE 2 | Cell line 1 | Tissue 2 | 62 |
| GENE 1 | Cell line 1 | Tissue 2 | 65 |
| GENE 2 | Cell line 2 | Tissue 2 | 70 |

For each perturbagen use average ranks (lower is better) from A to perform:
- Mann-Whitney U test for significance (with FDR correction)
- Estimate GREx antagonism pseudo z score: $-\frac{Hodges - Lehnman\ estimator_{compound}}{SD\ average\ ranks\ of\ all\ compounds}$

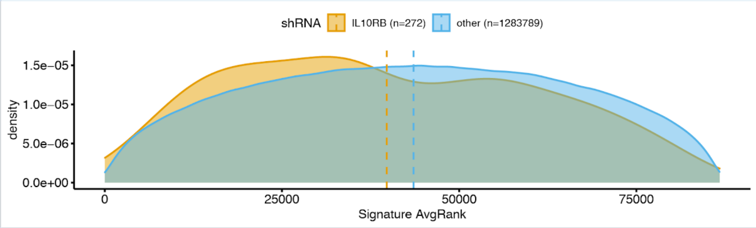


#### C. Gene prioritization approach (currenlty for shRNA; also CRISPR and overexpression in the future)

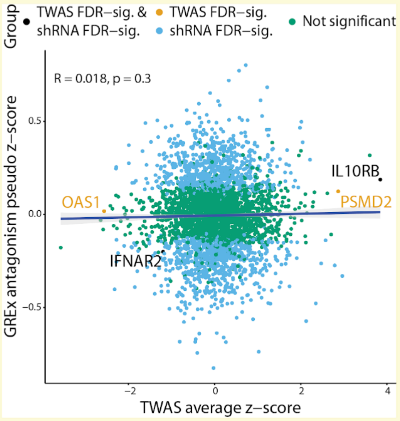


z.combined = GREx antagonism pseudo z-score + TWAS average z-score
Joint statistic (observed p value) is based on z.combined for prioritization

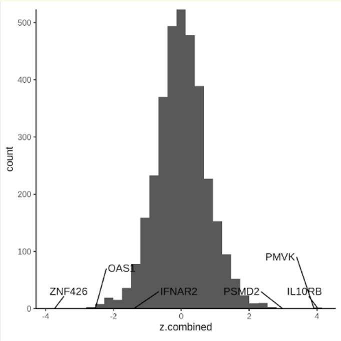


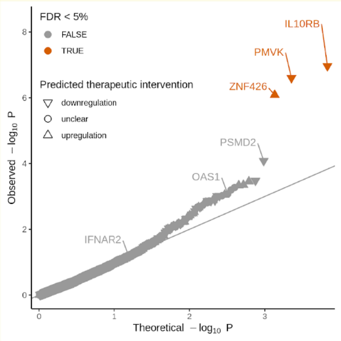

**Supplementary Figure 1. Gene target prioritization approach.** **Panel A.** Each signature from the perturbagen signature library (e.g. *IL10RB* shRNA treatment for 96 hours in MCF7 cells) is assessed for its ability to reverse the trait-associated imputed transcriptomes. **Panel B.** Signatures are grouped by peturbagen (either shRNA or compound) and we first test whether the signatures for a specific perturbagen are more likely to be ranked higher or lower (Mann-Whitney U test); then we obtain a GREx antagonism pseudo z-score as follows: $-\frac{Hodges{-}Lehmann\ estimator_{perturbagen}}{SD\ average\ ranks\ of\ all\ perturbagens}$(terms compound and peturbagen are used interchangeably). **Panel C.** Identification of candidate gene targets by integrating TWAS gene-trait associations and predicted effects of shRNAs in reversing COVID-19-associated transcriptomes. On the left (scatter plot), the x-axis corresponds to the average TWAS z-score ($z_{TWAS}$) across all EpiXcan tissues that have at least one FDR significant gene-trait association and the y-axis corresponds to the GREx antagonism pseudo z-score ($pseudo\ z_{GReX\ antagonism}$) which is defined as the negative Hodges-Lehmann estimator (of the median difference between that specific shRNA vs. all other shRNAs) divided by the standard deviation of the ranks of the compounds ( $-\frac{Hodges{-}Lehmann\ estimator_{perturbagen}}{SD\ average\ ranks\ of\ all\ perturbagens}$). A positive pseudo z-score is interpreted as a potentially therapeutic shRNA whereas a negative pseudo z-score would suggest that the shRNA is not antagonizing the imputed transcriptome and is thus likely to exacerbate the phenotype. Of note is that we only have shRNA gene expression signatures for 4,302 genes which is a subset of the genes that are reliably imputed from the TWAS. At the center, we see the histogram of the combined z scores ($z_{combined} = \overline{z_{TWAS}} + pseudo\ z_{GReX\ antagonism}$). On the right (QQ plot same as in Figure 2C), we show the p value corresponding to the joint statistic of the two approaches ( $z_{combined}$) described above against the null. FDR-significant candidate genes are labelled orange (whereas non FDR-significant are grey) and we also provide the direction of the predicted therapeutic intervention when this can be determined (upregulation or downregulation). *IL10RB*, *PMVK* and *ZNF426* are the three FDR-significant target genes, *PSMD2*, *OAS1* and *IFNAR2* are also displayed since they were FDR-significant TWAS genes to demonstrate the added value of the approach.

### Supplementary Figure 2

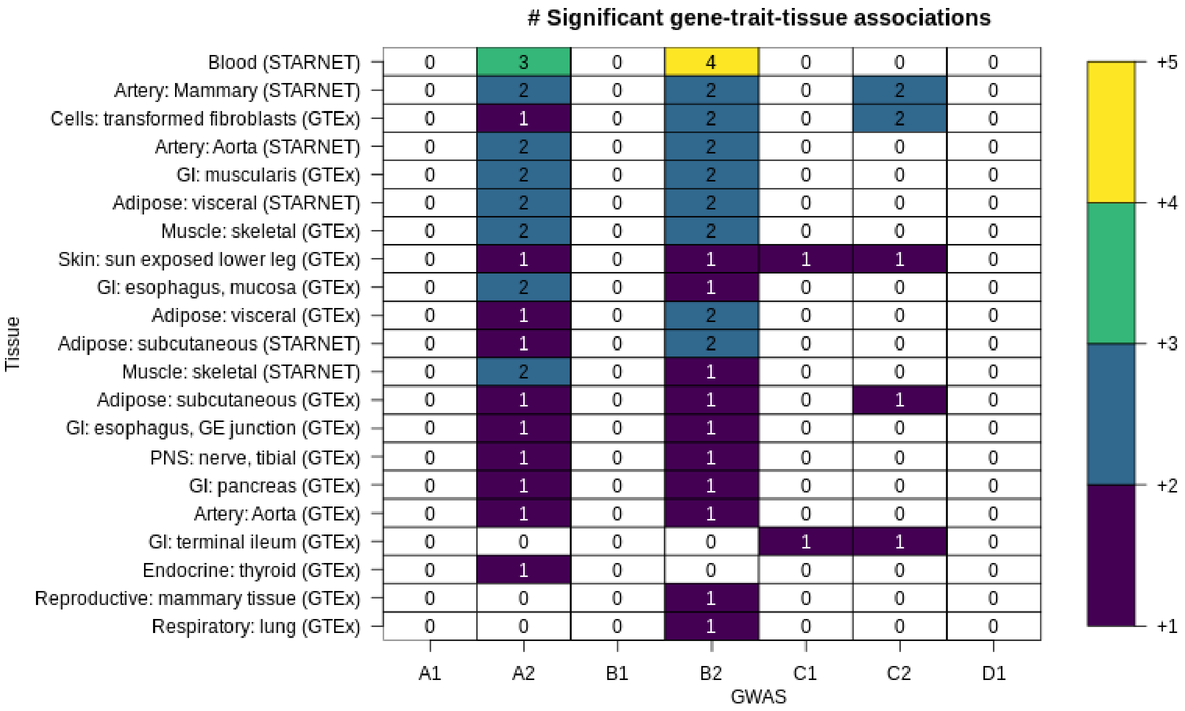


**<u>Supplementary Figure 2. TWAS gene-trait-tissue association counts per tissue and COVID-19 phenotype.</u>** Only FDR-significant associations are shown. To estimate FDR-adjusted p values (significant if FDR-adjusted p ≤ 0.05) we consider all phenotypes and tissues. A1: Very severe respiratory confirmed COVID vs. not hospitalized COVID ; A2: Very severe respiratory confirmed COVID vs. population; B1: Hospitalized COVID vs. not hospitalized COVID; B2: Hospitalized COVID vs. population; C1: COVID vs. lab/self-reported negative; C2: COVID vs. population; D1: predicted COVID from self-reported symptoms vs. predicted or self-reported non-COVID.

### Supplementary Figure 3

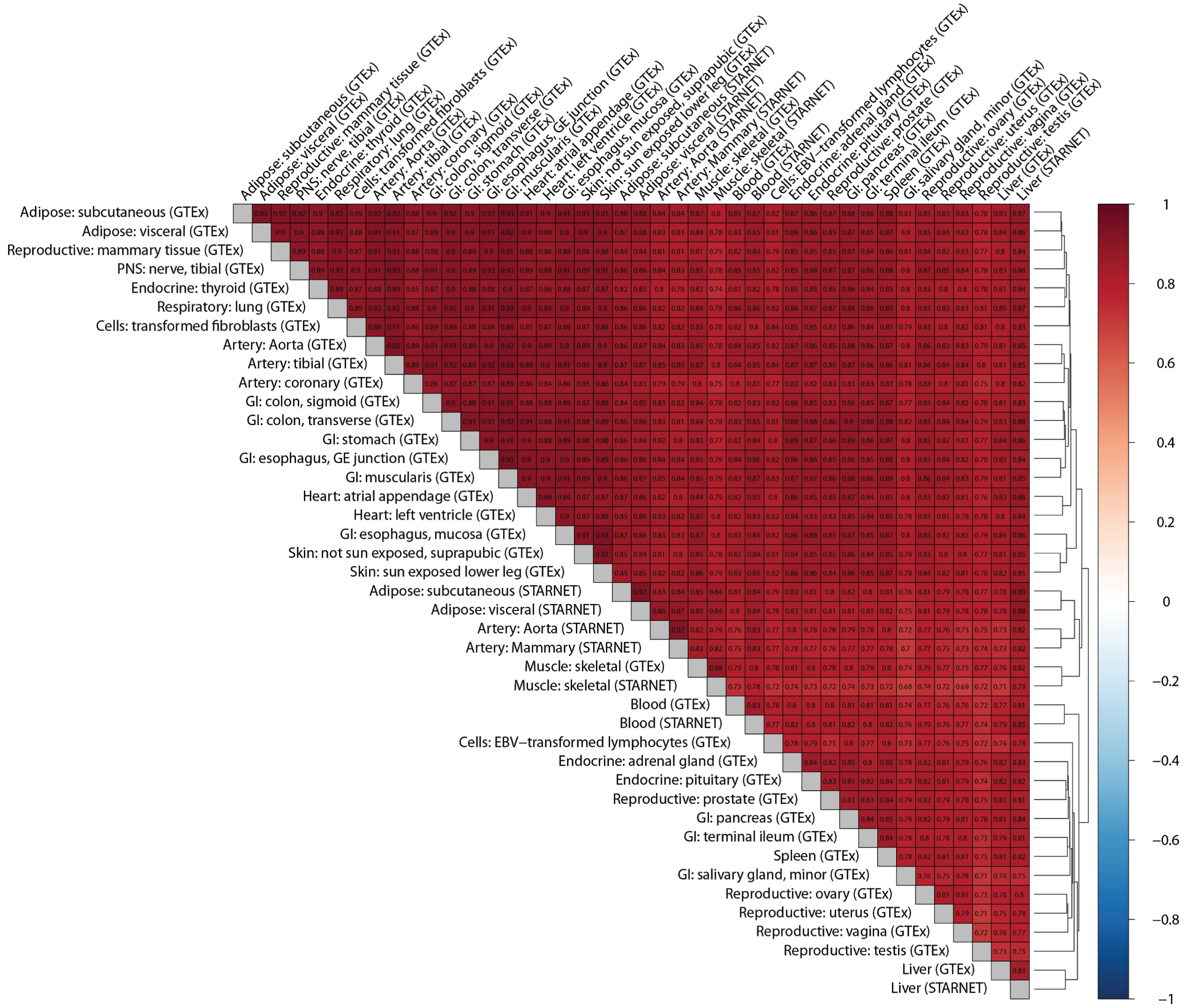


**<u>Supplementary Figure 3. Correlation of genetically regulated gene expression (GREx) across all tissues considering all COVID-19 phenotypes.</u>** Correlation is calculated for imputed expression changes with the Pearson method. Dendrogram on the right edge is shown from Ward hierarchical clustering.

#### Supplementary Figure 4

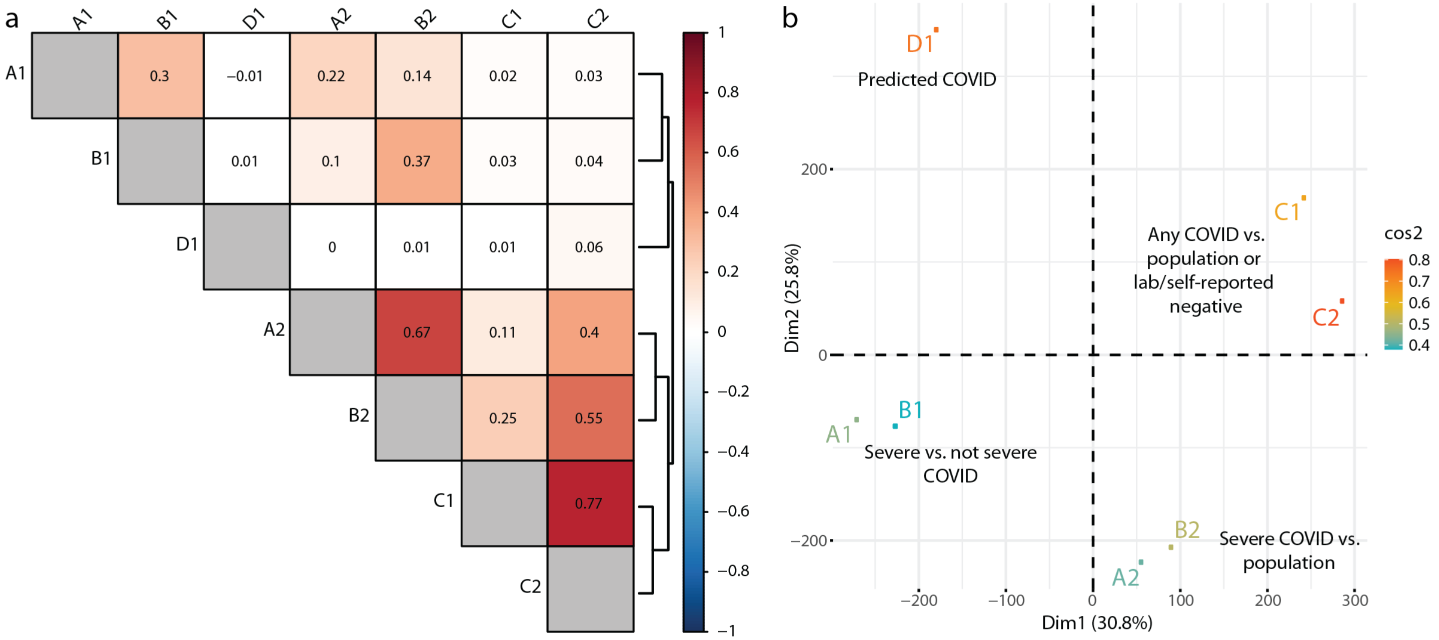


**Supplementary Figure 4. Panel a**. Correlation of GREx across COVID phenotypes taking into account all tissue models. Correlation is calculated for imputed expression changes with the Pearson method. Dendrogram on the right edge is shown from Ward hierarchical clustering. **Panel b.** PCA of GREx of COVID phenotypes showing clustering of phenotypes (e.g. A1&B1). The sums of the squared cosines of the first two principal components (PCs: Dim1 and Dim2) for each phenotype are color-coded as shown in the legend on the right and represent the importance of these PCs for each phenotype. A1: Very severe respiratory confirmed COVID vs. not hospitalized COVID ; A2: Very severe respiratory confirmed COVID vs. population; B1: Hospitalized COVID vs. not hospitalized COVID; B2: Hospitalized COVID vs. population; C1: COVID vs. lab/self-reported negative; C2: COVID vs. population; D1: predicted COVID from self-reported symptoms vs. predicted or self-reported non-COVID.

### Supplementary Figure 5

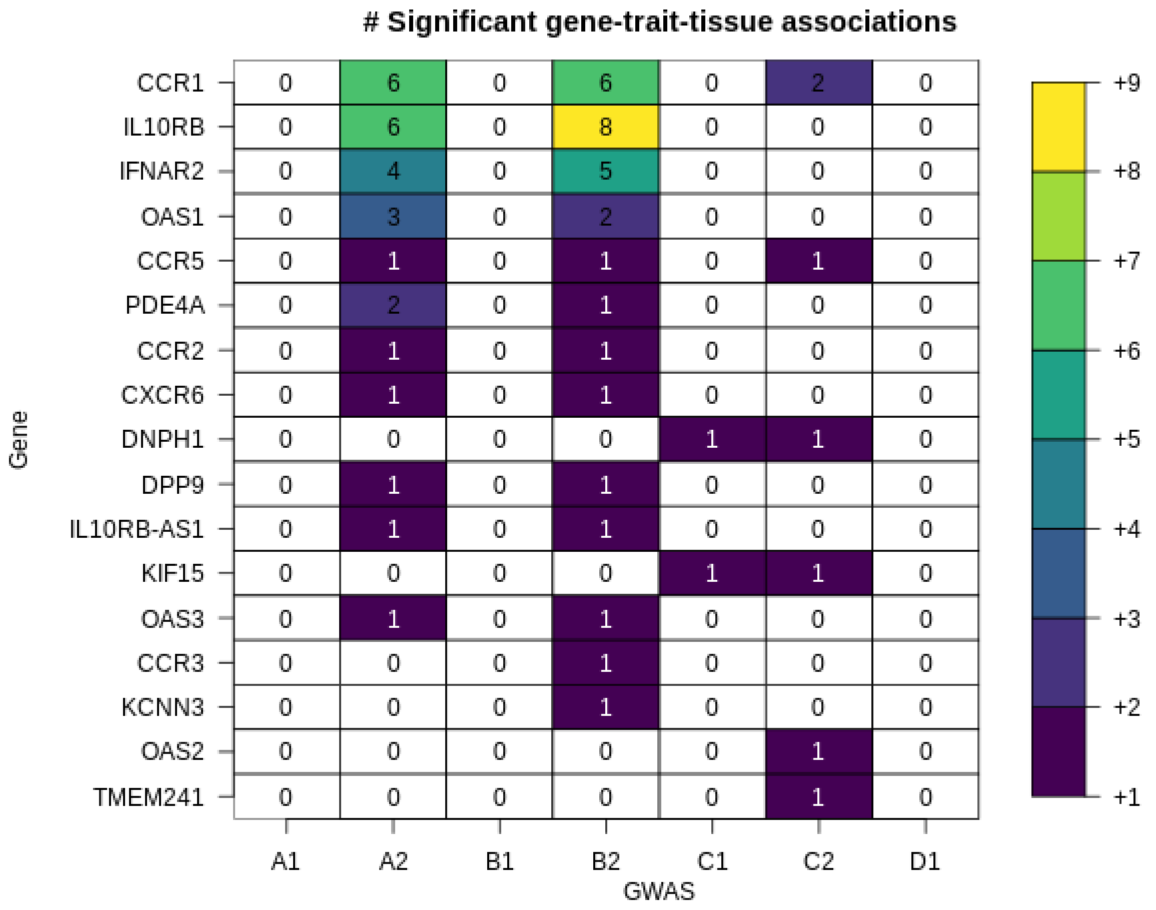


**<u>Supplementary Figure 5. TWAS gene-trait-tissue association counts per gene and COVID-19 phenotype (considering all tissue models).</u>** Only FDR-significant associations are shown. A1: Very severe respiratory confirmed COVID vs. not hospitalized COVID ; A2: Very severe respiratory confirmed COVID vs. population; B1: Hospitalized COVID vs. not hospitalized COVID; B2: Hospitalized COVID vs. population; C1: COVID vs. lab/self-reported negative; C2: COVID vs. population; D1: predicted COVID from self-reported symptoms vs. predicted or self-reported non-COVID.

### Supplementary Figure 6

#### A. OUTCOME: COVID-19 ASSOCIATED DEATH

### IL10RB

| HARE Population | IL10RB –> Death Beta | SE | n | g | 95% CI | weight |
|---|---|---|---|---|---|---|
| EUR | 0.12 | 0.0392 | 14262 | 0.12 | [ 0.04; 0.20] | 65.4% |
| AFR | 0.08 | 0.0646 | 5828 | 0.08 | [−0.05; 0.20] | 24.1% |
| HIS | 0.10 | 0.1003 | 2870 | 0.10 | [−0.10; 0.30] | 10.0% |
| ASN | 1.00 | 0.4644 | 266 | 1.00 | [ 0.09; 1.91] | 0.5% |
| Fixed effect model | | | 23226 | 0.11 | [ 0.05; 0.17] | 100.0% |
| Prediction interval | | | | | [−0.15; 0.37] | |

Heterogeneity: $I^2 = 24\%$, $\tau^2 = 0.0019$, $p = 0.27$

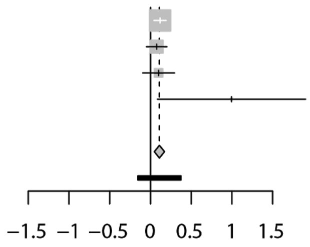


##### IFNAR2

| HARE Population | IFNAR2 –> Death Beta | SE | n | g | 95% CI | weight |
|---|---|---|---|---|---|---|
| EUR | −0.05 | 0.0392 | 14262 | −0.05 | [−0.13; 0.03] | 66.0% |
| AFR | 0.04 | 0.0659 | 5828 | 0.04 | [−0.09; 0.17] | 23.4% |
| HIS | −0.18 | 0.1005 | 2870 | −0.18 | [−0.38; 0.01] | 10.0% |
| ASN | −0.77 | 0.4214 | 266 | −0.77 | [−1.60; 0.05] | 0.6% |
| Fixed effect model | | | 23226 | −0.05 | [−0.11; 0.01] | 100.0% |
| Prediction interval | | | | | [−0.51; 0.38] | |

Heterogeneity: $I^2 = 53\%$, $\tau^2 = 0.0070$, $p = 0.09$

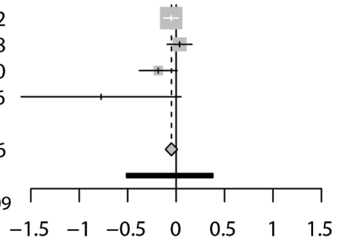


#### B. OUTCOME: COVID-19 SEVERITY

### IL10RB

| HARE Population | IL10RB –> Severity Beta | SE | n | g | 95% CI | weight |
|---|---|---|---|---|---|---|
| EUR | 0.06 | 0.0204 | 14262 | 0.06 | [ 0.02; 0.10] | 60.3% |
| AFR | 0.08 | 0.0297 | 5828 | 0.08 | [ 0.02; 0.14] | 28.4% |
| HIS | 0.06 | 0.0484 | 2870 | 0.06 | [−0.04; 0.15] | 10.7% |
| ASN | 0.46 | 0.2102 | 266 | 0.46 | [ 0.05; 0.88] | 0.6% |
| Fixed effect model | | | 23226 | 0.07 | [ 0.04; 0.10] | 100.0% |
| Prediction interval | | | | | [−0.05; 0.19] | |

Heterogeneity: $I^2 = 22\%$, $\tau^2 = 0.0004$, $p = 0.28$

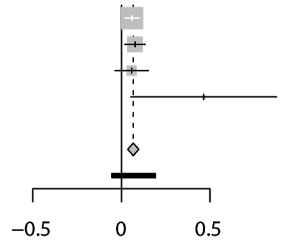


##### IFNAR2

| HARE Population | IFNAR2 –> Severity Beta | SE | n | g | 95% CI | weight |
|---|---|---|---|---|---|---|
| EUR | −0.06 | 0.0203 | 14262 | −0.06 | [−0.10; −0.02] | 60.3% |
| AFR | −0.08 | 0.0296 | 5828 | −0.08 | [−0.13; −0.02] | 28.4% |
| HIS | −0.09 | 0.0482 | 2870 | −0.09 | [−0.19; 0.00] | 10.7% |
| ASN | −0.41 | 0.2037 | 266 | −0.41 | [−0.81; −0.01] | 0.6% |
| Fixed effect model | | | 23226 | −0.07 | [−0.10; −0.04] | 100.0% |
| Prediction interval | | | | | [−0.15; 0.01] | |

Heterogeneity: $I^2 = 6\%$, $\tau^2 < 0.0001$, $p = 0.37$

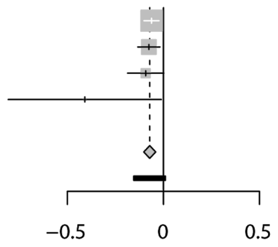


**Supplementary Figure 6. Transethnic meta-analysis for *IL10RB* and *IFNAR2* GReX with COVID-19 outcomes: death (A) and severity score (B).**

#### Supplementary Figure 7

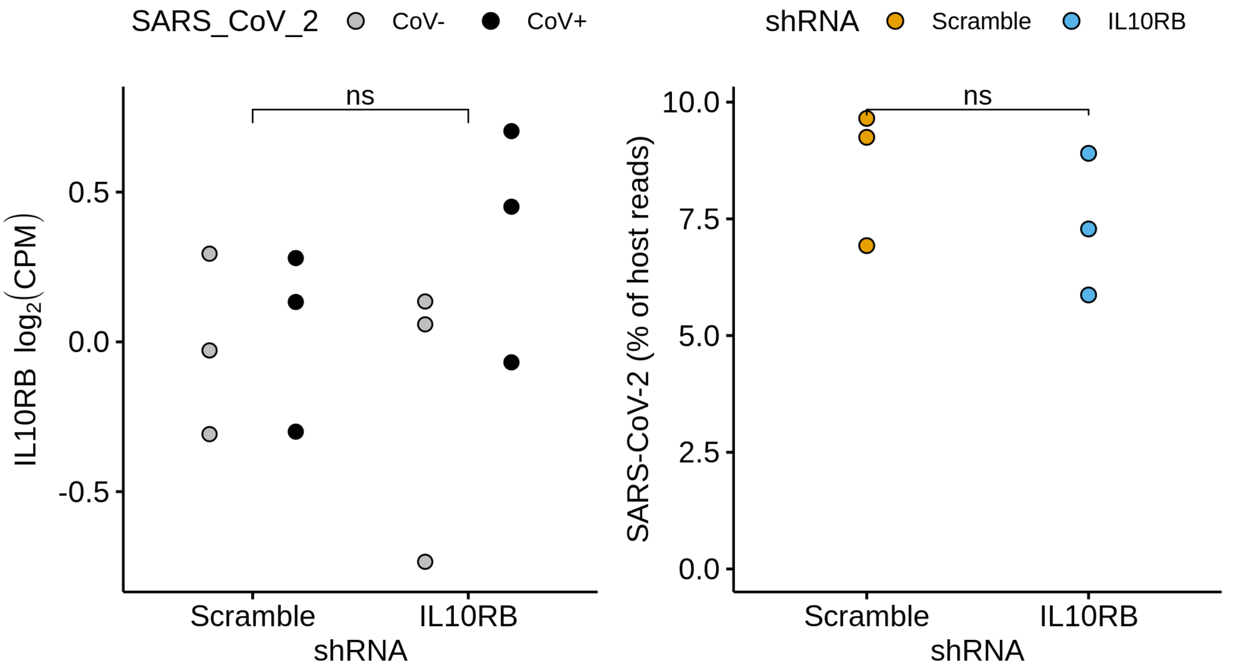


**Supplementary Figure 7. Effect of IL10RB shRNA on SARS-CoV-2 viral load in hiPSC-derived NGN-2 glutamatergic neurons.** shRNAs for IL10RB was used to knock-down *IL10RB* in hiPSC-derived NGN2-glutamatergic neurons. ***, ** and * correspond to p values from the linear model as ≤ 0.001, 0.01 and 0.05, respectively. For the SARS-CoV-2 viral load (right panel) we perform pairwise comparison with unpaired t-test; ***, ** and * correspond to p values of ≤ 0.001, 0.01 and 0.05, respectively.

Supplementary Figure 8

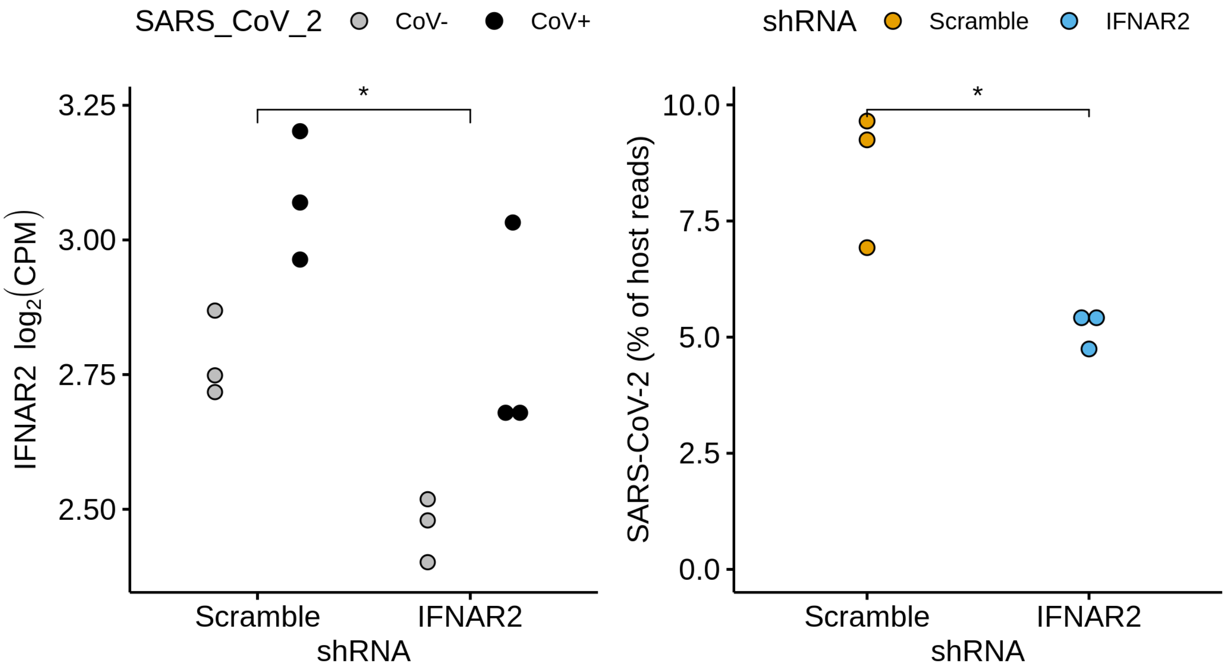


**Supplementary Figure 8. Effect of IFNAR2 shRNA on SARS-CoV-2 viral load in hiPSC-derived NGN-2 glutamatergic neurons.** shRNAs for IFNAR2 was used to knock-down *IFNAR2* in hiPSC-derived NGN2-glutamatergic neurons. ***, ** and * correspond to p values from the linear model of ≤ 0.001, 0.01 and 0.05, respectively. For the SARS-CoV-2 viral load (right panel) we perform pairwise comparison with unpaired t-test; ***, ** and * correspond to p values of ≤ 0.001, 0.01 and 0.05, respectively.

#### Supplementary Figure 9

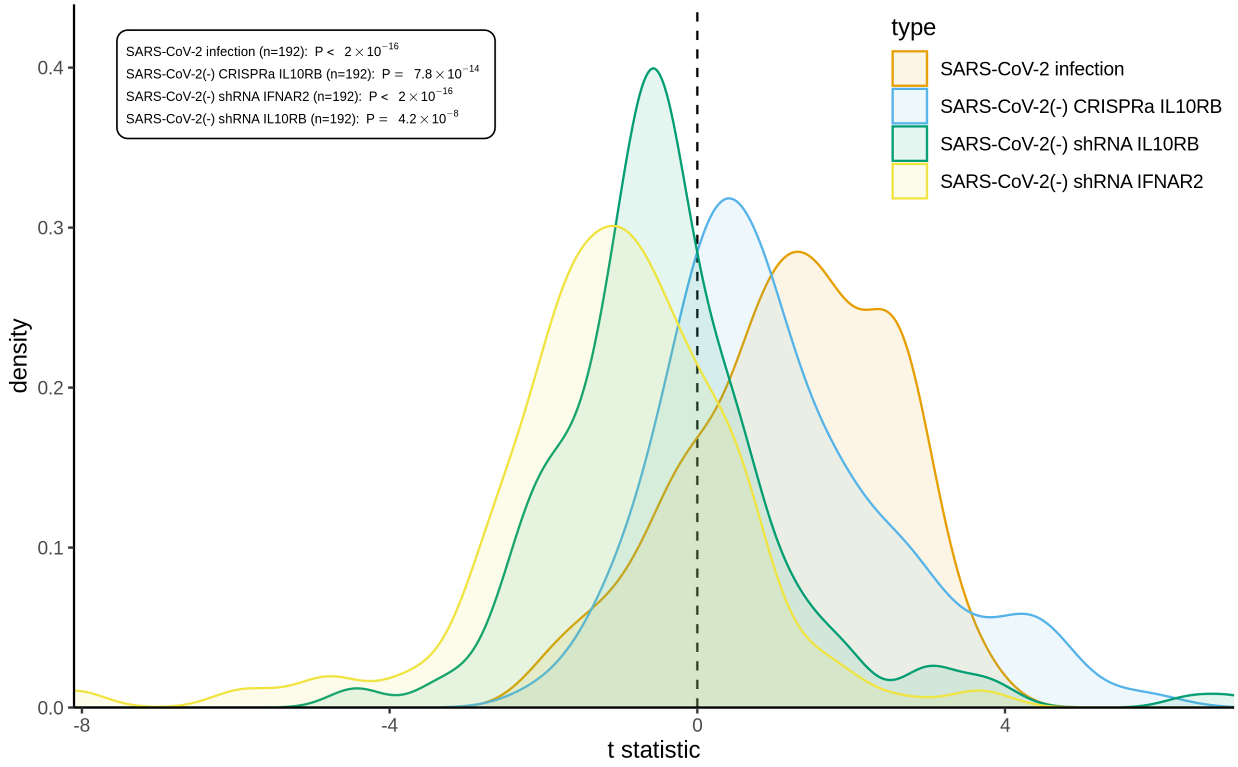


**Supplementary Figure 9. Competitive betacoronavirus gene set enrichment analysis in hiPSC-derived NGN-2 glutamatergic neurons.** Distribution of competitive enrichment t statistics for gene sets that correspond to betacoronavirus relevant gene sets e.g. infections across different cell systems and tissues (n=192; pruned betacoronavirus gene sets with a Jaccard index filter of 0.2). P values are from sign test against a theoretical median of 0.

### Supplementary Figure 10

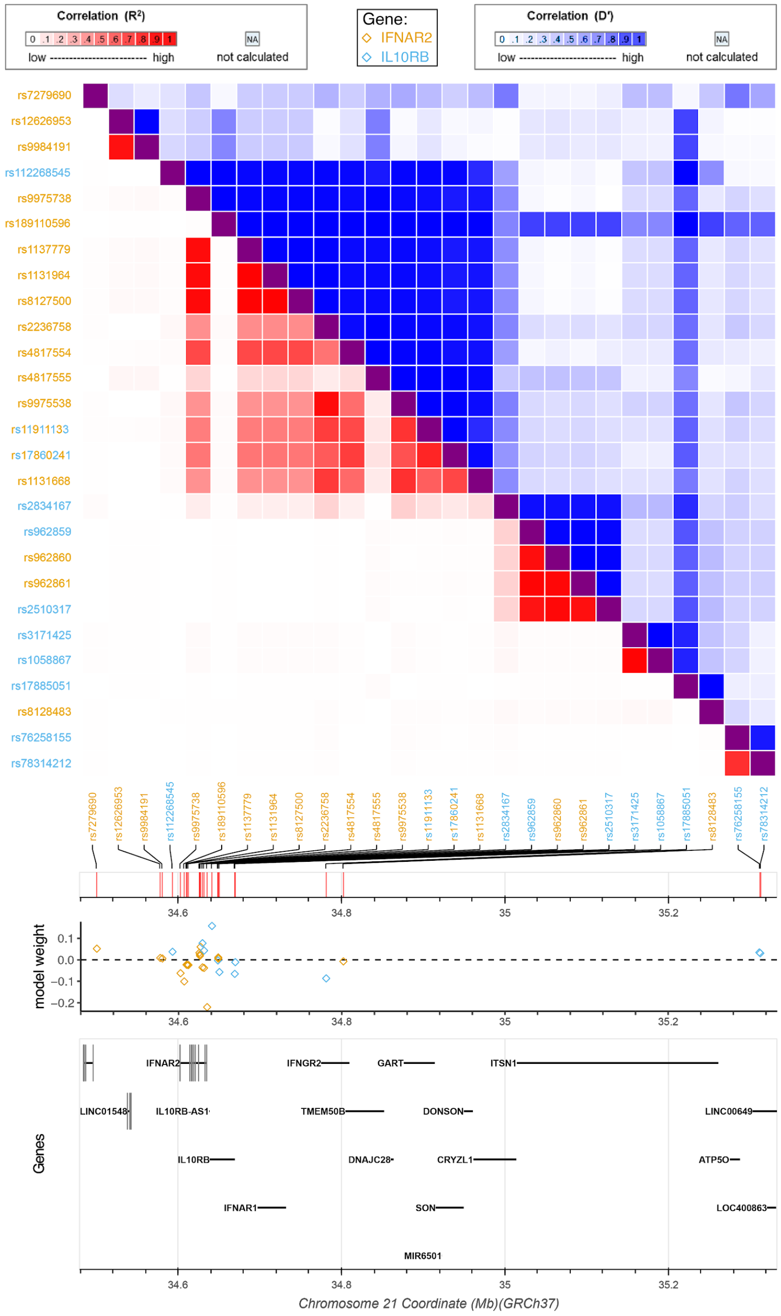


**<u>Supplementary Figure 10. Correlation map of SNPs with sizable contribution to the Blood (STARNET) models of *IFNAR2* and *IL10RB*.</u>** SNPs that are used for *IFNAR2* are orange, *IL10RB* are blue and those used by both have these two colors alternating by letter. Top panel is correlation ($R^2$ and D'), middle panel shows the model weights and bottom panel the genes in the region. Only SNPs with a model prior ≥ 2 for each model are shown; SNP correlation based on 1000G reference panel.

#### Supplementary Figure 11

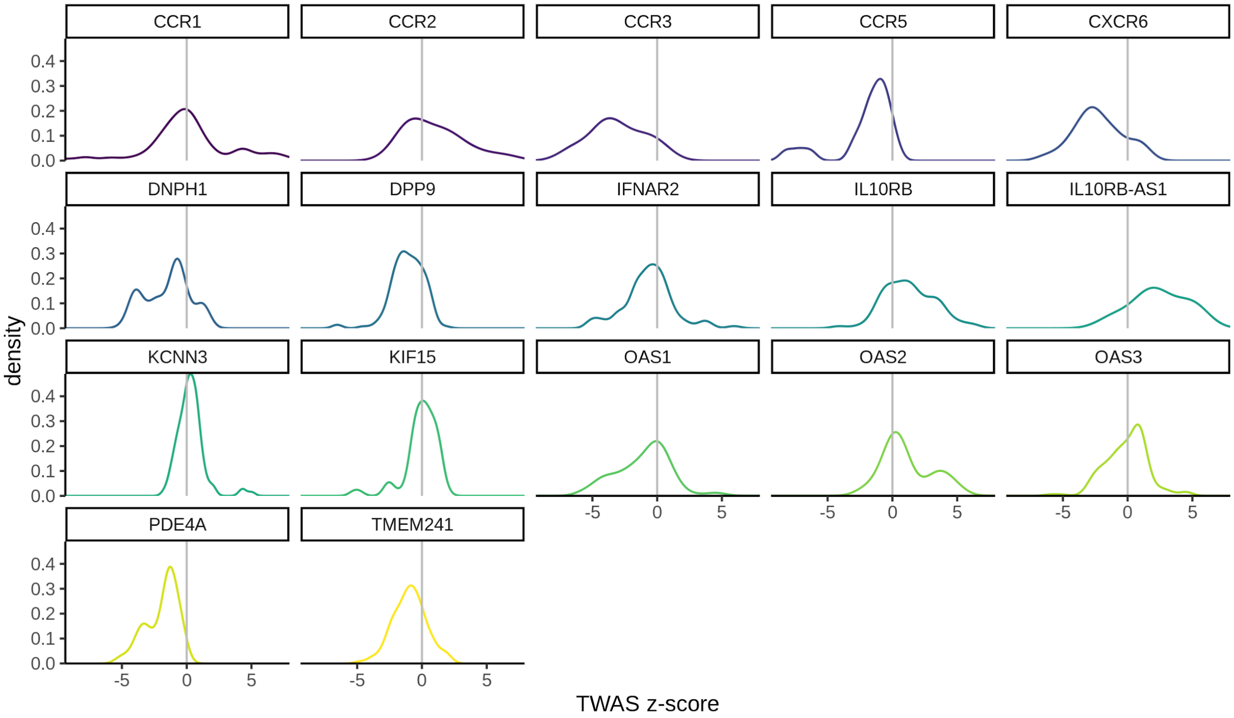


**Supplementary Figure 11. Density plots of TWAS association z-scores for FDR-significant genes (across all 7 COVID-19 phenotypes and 42 tissues).** For some genes such as *IL10RB* there is a relatively consistent shift of the z-scores to one direction (e.g. right) whereas in other genes such as *IFNAR2* there are both low and high z-score values suggesting phenotype and/or tissue specificity. FDR-significant genes (FDR-adjusted p≤0.05) for all COVID-19 phenotypes and tissues are displayed.

### Supplementary Figure 12

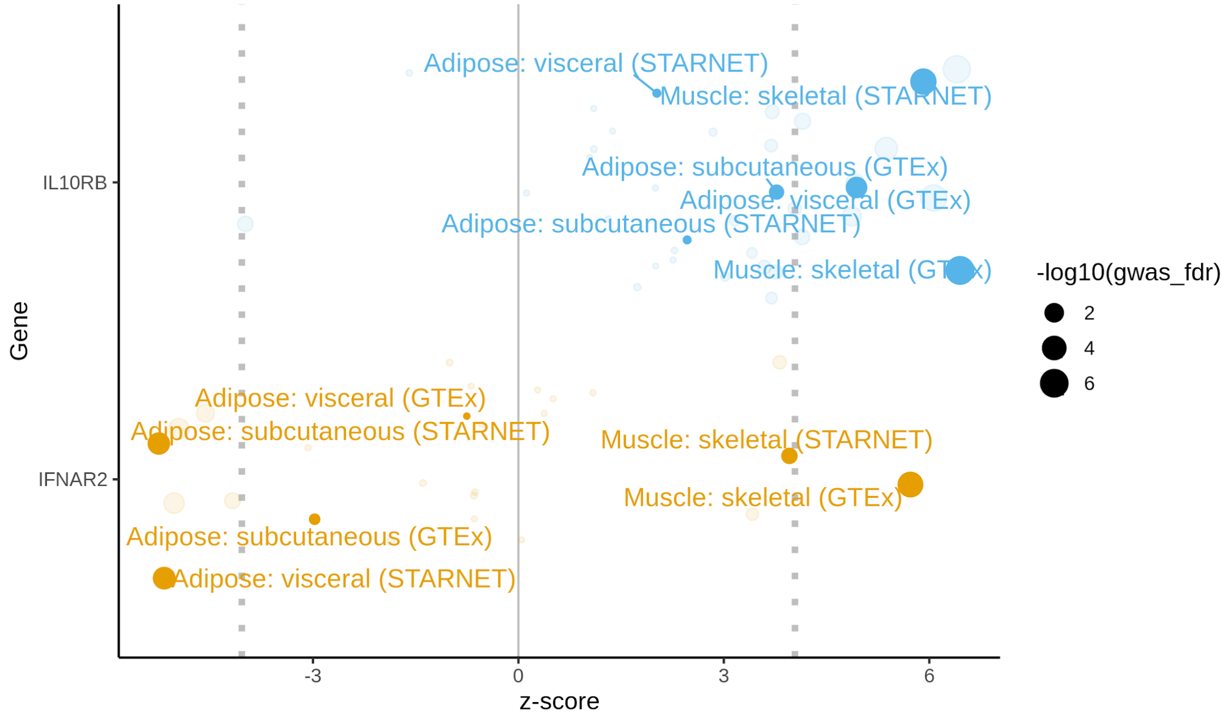


**<u>Supplementary Figure 12. Comparing tissue specificity for adipose and muscle tissues of *IL10RB* and *IFNAR2* TWAS z-scores.</u>** *IFNAR2* (blue) TWAS z-scores are consistently low for adipose tissue and high for skeletal muscle - this effect is consistent for related tissues (e.g. visceral and subcutaneous adipose tissue) and across cohorts (STARNET and GTEx). No such tissue specificity is observed in *IL10RB* (orange). Only the B2 phenotype for COVID-19 associated hospitalization is considered (FDR for B2 is displayed on the right) and FDR significance levels for z-scores are denoted with vertical dotted lines. Tissue z-scores not corresponding to adipose or skeletal muscle tissue are faded. Of note is that the faded blue dot for IL10RB that has a negative z-score that is close to significance corresponds to Cells: transformed fibroblasts (GTEx).

### SUPPLEMENTARY TABLES

#### Supplementary Table 1

| Category: tissue/cell (Cohort) | Transcriptomic imputation method used | Used in gene targeting and drug repurposing analysis? |
|---|---|---|
| Adipose: subcutaneous (GTEx) | EpiXcan | Yes |
| Adipose: subcutaneous (STARNET) | EpiXcan | Yes |
| Adipose: visceral (GTEx) | EpiXcan | Yes |
| Adipose: visceral (STARNET) | EpiXcan | Yes |
| Artery: Aorta (GTEx) | EpiXcan | Yes |
| Artery: Aorta (STARNET) | EpiXcan | Yes |
| Artery: coronary (GTEx) | EpiXcan | No |
| Artery: Mammary (STARNET) | EpiXcan | Yes |
| Artery: tibial (GTEx) | PrediXcan | No |
| Blood (GTEx) | EpiXcan | No |
| Blood (STARNET) | EpiXcan | Yes |
| Cells: EBV-transformed lymphocytes (GTEx) | PrediXcan | No |
| Cells: transformed fibroblasts (GTEx) | PrediXcan | No |
| Endocrine: adrenal gland (GTEx) | EpiXcan | No |
| Endocrine: pituitary (GTEx) | PrediXcan | No |
| Endocrine: thyroid (GTEx) | PrediXcan | No |
| GI: colon, sigmoid (GTEx) | EpiXcan | No |
| GI: colon, transverse (GTEx) | EpiXcan | No |
| GI: esophagus, GE junction (GTEx) | EpiXcan | Yes |
| GI: esophagus, mucosa (GTEx) | EpiXcan | Yes |
| GI: muscularis (GTEx) | EpiXcan | Yes |
| GI: pancreas (GTEx) | EpiXcan | Yes |
| GI: salivary gland, minor (GTEx) | PrediXcan | No |
| GI: stomach (GTEx) | EpiXcan | No |
| GI: terminal ileum (GTEx) | EpiXcan | No |
| Heart: atrial appendage (GTEx) | EpiXcan | No |
| Heart: left ventricle (GTEx) | EpiXcan | No |
| Liver (GTEx) | EpiXcan | No |
| Liver (STARNET) | EpiXcan | No |
| Muscle: skeletal (GTEx) | EpiXcan | Yes |
| Muscle: skeletal (STARNET) | EpiXcan | Yes |
| PNS: nerve, tibial (GTEx) | PrediXcan | No |
| Reproductive: mammary tissue (GTEx) | EpiXcan | Yes |
| Reproductive: ovary (GTEx) | EpiXcan | No |
| Reproductive: prostate (GTEx) | PrediXcan | No |
| Reproductive: testis (GTEx) | PrediXcan | No |
| Reproductive: uterus (GTEx) | PrediXcan | No |
| Reproductive: vagina (GTEx) | PrediXcan | No |
| Respiratory: lung (GTEx) | EpiXcan | Yes |
| Skin: not sun exposed, suprapubic (GTEx) | EpiXcan | No |

| | | |
|---|---|---|
| Skin: sun exposed lower leg (GTEx) | EpiXcan | Yes |
| Spleen (GTEx) | EpiXcan | No |

**Supplementary Table 1. The 42 transcriptomic imputation models used in this study.** Information is also provided about which imputation method was used and whether it was used for the gene targeting and drug repurposing pipelines.

#### Supplementary Table 2

| Short name | Phenotype | $n_{cases}$ | $n_{controls}$ | Ancestry superpopulation background | GWAS | TWAS results in |
|---|---|---|---|---|---|---|
| A1 | Very severe respiratory confirmed covid vs. not hospitalized covid | 269 | 688 | EUR | “A1_ALL” | Supplementary Appendix 3 |
| A2 | Very severe respiratory confirmed COVID vs. population | 4,336 | 623,902 | EUR + AMR | “A2_ALL_leave_23andme” | Supplementary Appendix 4 |
| B1 | Hospitalized COVID vs. not hospitalized COVID | 2,430 | 8,478 | ALL except EAS | “B1_ALL” | Supplementary Appendix 5 |
| B2 | Hospitalized COVID vs. population | 6,406 | 902,088 | EUR | “B2_ALL_eur_leave_23andme” | Supplementary Appendix 6 |
| C1 | COVID vs. lab/self-reported negative | 8,668 | 101,861 | ALL except EAS | “C1_ALL_leave_23andme” | Supplementary Appendix 7 |
| C2 | COVID vs. population | 14,134 | 1,284,876 | EUR | “C2_ALL_eur_leave_23andme” | Supplementary Appendix 8 |
| D1 | predicted COVID from self-reported symptoms vs. predicted or self-reported non-COVID | 3,204 | 35,728 | EUR | “D1_ALL” | Supplementary Appendix 9 |

**Supplementary Table 2. Overview of the GWAS summary statistics that were used.** Short name: short name of the phenotype. Phenotype: description of the phenotype. $n_{cases}$ and $n_{controls}$ correspond to the number of cases and controls taken into account for this study. Ancestry superpopulation background: ancestry superpopulations that were included in the GWAS. GWAS: GWAS summary statistics used. TWAS results in: Supplementary appendix where the TWAS results can be found. EUR, AMR, EAS stand for European, admixed American and East Asian ancestries. All refers to all the superpopulations as defined by the 1000 genomes project which includes the above plus African and South Asian ancestries.

### Supplementary Table 3

| Severity | Description | $n_{EUR}$ | $n_{AFR}$ | $n_{HIS}$ | $n_{ASN}$ |
|---|---|---|---|---|---|
| Mild | SARS-CoV-2+ | 10,851 | 4,113 | 2,221 | 217 |
| Moderate | SARS-CoV-2+ and hospitalized with or without low flow oxygen therapy | 2,301 | 1,187 | 439 | 26 |
| Severe | SARS-CoV-2+ and hospitalized with either ventilation, intubation, extracorporeal membrane oxygenation (EMCO), dialysis vasopressors or high flow oxygen therapy | 383 | 266 | 93 | 8 |
| Death | COVID-19 related death | 727 | 262 | 117 | 15 |

**Supplementary Table 3. COVID severity scale developed by VINCI and the MVP COVID-19 Science Initiative.** Description and counts for each HARE-based population are provided.

#### Supplementary Table 4

| Severity | Description |
|---|---|
| Control | No COVID-19 |
| Moderate | COVID-19 with abnormal (<94%) O2 saturation or pneumonia on imaging |
| Severe | COVID-19 with use of high-flow nasal cannula (HFNC), non-rebreather mask (NRB), bilevel positive airway pressure (BIPAP) or mechanical ventilation and no vasopressor use, and based on CrCl greater than 30 and alanine aminotransferase (ALT) less than 5× the upper limit of normal. |
| Severe with end-organ damage | COVID-19 as Severe but with use of vasopressors, or based on CrCl less than 30, new renal replacement therapy (hemodialysis/continuous veno-venous hemofiltration) or ALT more than 5× the upper limit of normal |

**Supplementary Table 4. COVID severity scale developed by the Mount Sinai COVID-19 Biobank.** Severity score has been previously characterized in detail[4].

### Supplementary Table 5

| Rank | Compound | Compound MW FDR | Mechanism of action (MOA) | MOA Rank | Experimental/epidemiological support (XP) or speculation/ hypothesis/ poorly controlled studies (HY) | Notes and challenges |
|---|---|---|---|---|---|---|
| 1 | **<u>imiquimod</u>** | 0.0001 | interferon inducer\|toll-like receptor agonist | | HY[19–21] | Topical use but systemic absorption [22] |
| 2 | **nelfinavir** | 0.005 | HIV protease inhibitor | 1 | XP[23,24] | No longer recommended since 2019 |
| 3 | **saquinavir** | 0.03 | HIV protease inhibitor | 1 | XP[23] | |
| 4 | **<u>everolimus</u>** | $4.6 \times 10^{-9}$ | mTOR inhibitor | 20 | HY[25] | Non-formulary (VA) |
| 5 | **<u>azathioprine</u>** | $0.0008 \times 10^{-4}$ | dehydrogenase inhibitor | | HY[26] | |
| 6 | **nisoldipine** | 0.02 | calcium channel blocker | 33 | | Non-formulary (VA) |
| 7 | cerulenin | $3.6 \times 10^{-11}$ | fatty acid synthase inhibitor | | XP: Similar MOA[27] | <u>Excluded:</u> not FDA approved |
| 8 | pyrvinium-pamoate | $5.1 \times 10^{-20}$ | androgen receptor antagonist | 7 | Other CoV XP[28] | <u>Excluded:</u> discontinued |
| 9 | **<u>retinol</u>** | 0.04 | retinoid receptor ligand | | HY[29] | Vitamin A |
| 10 | selamectin | 0.01 | nematocide | | | <u>Excluded:</u> not approved for human use |

**<u>Supplementary Table 5. Top 10 readily available candidate compounds.</u>** Ranking of the compound is provided while considering only readily available compounds (bold if FDA-approved, underlined if well-powered for population-level analysis). Compound MW FDR: FDR-adjusted p-values for the Mann-Whitney U test while considering compounds from all clinical phases (not just "launched"). MOA Rank: rank of the medication mechanism of action among "launched" compounds. All literature support lacks any medical evidence for use in clinical practice. For the whole table please refer to Supplementary Appendix 11.

### Supplementary Table 6

| Compound | Indication ICD-9 | Indication name | Abbreviation |
|---|---|---|---|
| azathioprine | 571.42 | Autoimmune hepatitis | AIHP |
| | 555.0,555.1,555.2,555.9 | Crohn’s disease | CRHN |
| | 710.3, 710.4 | Dermatomyositis/polymyositis | DERM |
| | 695.10,695.11,69.12,695.19 | Erythema multiforme | ERYT |
| | 996.83,V42.1,V42.2 | Heart transplant | HRTR |
| | 287.31 | Idiopathic thrombocytopenic purpura | IDTP |
| | 996.81,V42.0 | Kidney transplant | KDTR |
| | 996.82,V42.7 | Liver transplant | LVTR |
| | 996.84,V42.6 | Lung transplant | LUTR |
| | 358.00,358.01 | Myasthenia gravis | MYAG |
| | 583.81 | Nephritis and nephropathy, not specified as acute or chronic, in diseases classified elsewhere | NEPH |
| | 391.0,393,420.0,420.90,420.91,420.99,423.1,423.2 | Non-infectious pericarditis | NPER |
| | 694.4 | Pemphigus | PEMP |
| | 696.0,696.1,696.8 | Psoriasis (with arthropathy and other) | PSOR |
| | 714.0,714.1,714.2,714.30,714.31,714.32,714.33,714.4,714.8,714.81,714.89,714.9 | Rheumatoid arthritis | RHEU |
| | 556.0,556.1,556.2,556.3,556.4,556.5,556.6,556.8,556.9 | Ulcerative colitis | ULCR |
| | 360.11,360.12,363.01,363.03,363.04,363.05,363.06,363.07,363.08,363.10,363.11,363.12,363.13,363.14,363.15,363.20,363.21,363.22,364.00,364.01,364.02,364.03,364.04,364.05,364.10,364.11,364.21,364.22,364.23,364.24 | Uveitis | UVET |
| | 446.4 | Wegener’s granulomatosis | WEGR |
| everolimus | 209.00,209.01,209.02,209.03,209.10,209.11,209.12,209.13,209.14,209.15,209.16,209.17,209.20,209.21,209.22,209.23,209.24,209.25,209.26,209.27,209.29,209.40,209.41,209.4 | Carcinoid tumors | CARC |

| | | | |
|---|---|---|---|
| | 2,209.43,209.50,209.51,209.52,209.53,209.54,209.55,209.56,209.57,209.60,209.61,209.62,209.63,209.64,209.65,209.66,209.67,209.69 | | |
| | 996.83,V42.1,V42.2 | Heart transplant | HRTR |
| | 201.00,201.01,201.02,201.03,201.04,201.05,201.06,201.07,201.08,201.10,201.11,201.12,201.13,201.14,201.15,201.16,201.17,201.18,201.20,201.21,201.22,201.23,201.24,201.25,201.26,201.27,201.28,201.40,201.41,201.42,201.43,201.44,201.45,201.46,201.47,201.48,201.50,201.51,201.52,201.53,201.54,201.55,201.56,201.57,201.58,201.60,201.61,201.62,201.63,201.64,201.65,201.66,201.67,201.68,201.70,201.71,201.72,201.73,201.74,201.75,201.76,201.77,201.78,201.90,201.91,201.92,201.93,201.94,201.95,201.96,201.97,201.98,V10.72 | Hodgkin's disease | HODG |
| | 996.81,V42.0 | Kidney transplant | KDTR |
| | 996.82,V42.7 | Liver transplant | LVTR |
| | 996.84,V42.6 | Lung transplant | LUTR |
| | 273.3 | Macroglobulinemia | MACR |
| | 189.0,198.0,V10.52 | Neoplasm of the kidney | NPKD |
| | 164.0,212.6 | Neoplasm of the thymus | NPTY |
| | 174.0,174.1,174.2,174.3,174.4,174.5,174.6,174.8,174.9,175.0,175.9,233.0,V10.3 | Non-benign Neoplasm of the breast (male and female) | NPBR |
| | 759.5 | Tuberous sclerosis | TUSC |
| imiquimod | 702.0 | Actinic keratosis | ACKE |
| | 173.01,173.11,173.21,173.31,173.41,173.51,173.61,173.71,173.81,173.91 | Basal cell carcinoma | BCCA |
| | 054.0,054.10,054.11,054.12,054.13,054.19,054.2,054.40,054.41, 054.42, 054.43, 054.44,054.49,054.5,054.6,054.7,054.71,054.72,054.73,054.74,054.79,054.8,054.9,058.10,058.11,058.12,058.21,058.29,058.81, | Herpes simplex virus (HSV) infection | HSVI |

| | | | |
|---|---|---|---|
| | 058.82,058.89 | | |
| | 078.10,078.11,078.12,078.19,079.4 | Viral warts & HPV | VHPV |
| nelfinavir | 042,079.53,V08 | HIV infection | HIVI |
| nisoldipine | 401.0,401.1,401.9,405.01,405.09,405.11,405.19,405.91,405.99 | Hypertension | HYPT |
| retinol | 264.0,264.1,264.2,264.3,264.4,264.5,264.6,264.7,264.8,264.9 | Vitamin A deficiency | VADE |
| saquinavir | 042,079.53,V08 | HIV infection | HIVI |

**Supplementary Table 6. FDA approved and off-label indications for medications in our analysis used as covariates.** For each compound, the indication ICD-9 codes (comma separated), name and abbreviation used in the models (Supplementary Appendix 12) are provided.

#### Supplementary Table 7

| | Severity Analysis Cohort | | | | | PheWAS Cohort |
|---|---|---|---|---|---|---|
| | EUR | AFR | HIS | ASN | ALL | EUR |
| Sample Size (%) | 14,262 (61.4%) | 5,828 (25.1%) | 2,870 (12.4%) | 266 (1.1%) | 23,226 (100%) | 296,407 (N/A) |
| Median Age (IQR) | 71 (16) | 64 (17) | 60 (27) | 50 (27) | 68 (18) | 71 (14) |
| Female (%) | 1,231 (8.6%) | 824 (14.1%) | 295 (10.3%) | 26 (9.8%) | 2,376 (10.2%) | 21,084 (7.1%) |
| Median Elixhauser (2yr) (IQR) | 4 (13) | 5 (14) | 0 (8) | 0 (4) | 4 (13) | N/A |
| COVID Severity<br>- Mild (%)<br>- Median (%)<br>- Severe (%)<br>- Death (%) | <br>10,851 (76.1%)<br>2,301 (16.1%)<br>383 (2.7%)<br>727 (5.1%) | <br>4,113 (70.6%)<br>1,187 (20.4%)<br>266 (4.6%)<br>262 (4.5%) | <br>2,221 (77.4%)<br>439 (15.3%)<br>93 (3.2%)<br>117 (4.1%) | <br>217 (81.6%)<br>26 (9.8%)<br>8 (3.0%)<br>15 (5.6%) | <br>17,402 (74.9%)<br>3,953 (17.0%)<br>750 (3.2%)<br>1,121 (4.8%) | N/A |
| Median ICD Count (IQR) | N/A | N/A | N/A | N/A | N/A | 136 (175) |
| Median length of record (IQR) | N/A | N/A | N/A | N/A | N/A | 4,575.4 (3,350.0) |

**Supplementary Table 7. Demographic characteristics of the MVP cohorts used in the GReX association with COVID-19 severity and death, and PheWAS.**

### Supplementary Table 8

| Gene | Population | n | Beta | SE | P | Bonferroni-adjusted p |
|---|---|---|---|---|---|---|
| IL10RB | ALL | 23226 | 0.067 | 0.016 | **$2.4\times10^{-05}$** | **$9.8\times10^{-05}$** |
| | EUR | 14262 | 0.060 | 0.020 | **$3.3\times10^{-03}$** | **$1.3\times10^{-02}$** |
| | AFR | 5828 | 0.077 | 0.030 | **$9.6\times10^{-03}$** | **$3.8\times10^{-02}$** |
| | HIS | 2870 | 0.058 | 0.048 | $2.3\times10^{-01}$ | $9.2\times10^{-01}$ |
| | ASN | 266 | 0.465 | 0.210 | **$2.7\times10^{-02}$** | $1.1\times10^{-01}$ |
| IFNAR2 | ALL | 23226 | -0.071 | 0.016 | **$6.2\times10^{-06}$** | **$2.5\times10^{-05}$** |
| | EUR | 14262 | -0.062 | 0.020 | **$2.3\times10^{-03}$** | **$9.0\times10^{-03}$** |
| | AFR | 5828 | -0.076 | 0.030 | **$1.0\times10^{-02}$** | **$4.1\times10^{-02}$** |
| | HIS | 2870 | -0.092 | 0.048 | $5.5\times10^{-02}$ | $2.2\times10^{-01}$ |
| | ASN | 266 | -0.409 | 0.204 | **$4.4\times10^{-02}$** | $1.8\times10^{-01}$ |

**Supplementary Table 8. GReX association with COVID-19 severity.** Bonferroni-adjustment is performed for $n_{genes} \times n_{outcomes} = 4$ for each population cohort.

### Supplementary Table 9

| Population / ethnicity | Sample Size (%) | Median Age (IQR) | Female (%) |
|---|---|---|---|
| White / Hispanic or Latino | 52 (9.2%) | 66 (21.25) | 24 (46.2%) |
| Black or African American / Hispanic or Latino | 10 (1.8%) | 52 (32) | 7 (70%) |
| Unknown / Hispanic or Latino | 107 (18.8%) | 64 (25) | 41 (38.3%) |
| More Than One Race / Hispanic or Latino | 15 (2.6%) | 64 (18) | 7 (46.7%) |
| American Indian or Alaska Native / Hispanic or Latino | 4 (0.7%) | 63.5 (17.5) | 1 (25%) |
| White / not Hispanic or Latino | 128 (22.5%) | 68 (28.5) | 54 (42.2%) |
| Black or African American / not Hispanic or Latino | 106 (18.7%) | 63 (16.5) | 54 (50.9%) |
| Unknown / not Hispanic or Latino | 12 (2.1%) | 62 (14.5) | 1 (8.3%) |
| More Than One Race / not Hispanic or Latino | 9 (1.6%) | 56 (12) | 4 (44.4%) |
| Asian / not Hispanic or Latino | 36 (6.3%) | 60 (22.25) | 11 (30.6%) |
| American Indian or Alaska Native / not Hispanic or Latino | 3 (0.5%) | 62 (7.5) | 0 (0%) |
| Native Hawaiian or Other Pacific Islander / not Hispanic or Latino | 1 (0.2%) | 68 (0) | 1 (100%) |
| White / unknown | 3 (0.5%) | 88 (4.5) | 2 (66.7%) |
| Unknown unknown | 82 (14.4%) | 62 (21.5) | 35 (42.7%) |

**Supplementary Table 9. Demographic characteristics of the Mount Sinai COVID-19 Biobank.**

#### Supplementary Table 10

| | Count (%) |
|---|---|
| # Individuals with number of samples below: | |
| - 1 | 176 (31.0%) |
| - 2 | 156 (27.5%) |
| - 3 | 110 (19.4%) |
| - 4 | 71 (12.5%) |
| - 5 | 39 (6.9%) |
| - 6 | 15 (2.6%) |
| - 7 | 1 (0.2%) |
| # of samples with COVID Severity below: | |
| - Control | 122 (10.1%) |
| - Moderate COVID-19 | 600 (49.6%) |
| - Severe COVID-19 | 269 (22.2%) |
| - Severe COVID-19 with EOD | 218 (18.0%) |

**Supplementary Table 10. Sample characteristics of the Mount Sinai COVID-19 Biobank.** The differential gene expression analysis is based on samples not individuals. Here we provide information about how many samples were taken from each individual and a breakdown by severity for the samples.

### Supplementary Table 11

| gRNA | Target gene | logFC | t | P value |
|---|---|---|---|---|
| IL10RB-1 | IL10RB | 2.67 | 5.67 | 0.0000009 |
| IL10RB-2 | IL10RB | 1.46 | 3.44 | 0.0012 |
| IL10RB-3 | IL10RB | 1.67 | 3.90 | 0.0003 |
| IL10RB-4 | IL10RB | 0.98 | 3.64 | 0.0007 |

**Supplementary Table 11. Effect of CRISPRa gRNAs on target gene.** Metrics are provided against scramble gRNA.

### Supplementary Table 12

| shRNA | Target gene | logFC | t | P value |
|---|---|---|---|---|
| IL10RB | IL10RB | 0.06 | 0.27 | 0.78 |
| IFNAR2 | IFNAR2 | -0.41 | -2.39 | 0.02 |

**Supplementary Table 12. Effect of shRNAs on target genes.** Metrics are provided against scramble shRNA

#### Supplementary Table 13

| Treatment | Target gene | logFC | t | P value |
|---|---|---|---|---|
| SARS-CoV-2 infection | IL10RB | 0.025 | 0.13 | 0.89 |
| SARS-CoV-2 infection | IFNAR2 | 0.3 | 3.70 | 0.00098 |

**Supplementary Table 13. Effect of SARS-CoV-2 infection on target genes (IL10RB and IFNAR2).** Metrics are provided against non SARS-CoV-2 infected cells taking into account (scramble gRNA and shRNA treatments).

### Supplementary Table 14

**Unique patient counts**

| | Ever prescribed[a] | Within 90 days prior to 2/15/2020[b] | Tested for SARS-CoV-2[c] | SARS-CoV-2+[d] |
|---|---|---|---|---|
| **Top 7 compounds** | 176,782 | 14,329 | 4,416 | 369 |
| imiquimod | 91,671 | 4,859 | 883 | 88 |
| everolimus | 921 | 350 | 105 | 7 |
| azathioprine | 21,180 | 5,765 | 1,419 | 123 |
| retinol | 62,564 | 3,362 | 2,018 | 152 |
| nelfinavir | 3,294 | 12 | 4 | 0 |
| saquinavir | 1,176 | 5 | 3 | 0 |
| nisoldipine | 47 | 7 | 0 | 0 |
| **Antiretrovirals[e]** | 30,102 | 24,269 | 6,693 | 777 |
| Non-protease inhibitors | 30,063 | 24,232 | 6,676 | 777 |
| NRTI | 29,888 | 22,866 | 6,237 | 722 |
| NNRTI | 19,192 | 4,913 | 1,216 | 121 |
| Other | 21,648 | 14,300 | 3,824 | 453 |
| Protease inhibitors (PI) | 15,519 | 3,826 | 1,101 | 114 |
| **Groups of patients diagnosed with HIV (ICD-9 042 or ICD-10 B20)** | | | | |
| A: No anti-HIV medications | 29,777 | 34,823 | 4,720 | 1,003 |
| B: Any of anti-HIV medications AND PI | 15,480 | 3,789 | 1,084 | 114 |
| C: Any of anti-HIV medications AND no PI | 14,583 | 20,443 | 5,592 | 663 |

**Supplementary Table 14. Unique patient counts for the VHA population-level analysis.** [a]Data through 12/31/2020. [b]Any prescription in the 90 days prior to 2/15/2020. [c]Any prescription in the 90 days prior to the date of first COVID-19 testing. [d]Any prescription in the 90 days prior to the first positive test for SARS-CoV-2 as of 11/30/2020. [e]Limited to patients with any HIV diagnosis codes (ICD-9 042 or ICD-10 B20) prior to the index date. Includes records for Veterans alive as of 2/15/2020.

### Supplementary Table 15

| Protease inhibitor | Patient count |
|---|---|
| **Darunavir** | 3,005 |
| **Ritonavir** | 1,651 |
| **Atazanavir** | 485 |
| **Lopinavir** | 142 |
| **Fosamprenavir** | 25 |
| **Nelfinavir** | 11 |
| **Tipranavir** | 6 |
| **Saquinavir** | 5 |
| **Indinavir** | 5 |

**Supplementary Table 15: Patients receiving prescriptions of specific anti-HIV protease inhibitor medications in the 90 day window prior to the SARS-CoV-2 test.** Counts are limited to 3,862 patients diagnosed with HIV and taking antiretroviral medications.

#### Supplementary Table 16

| gRNA target | Oligo Sequences forward | Oligo Sequences reverse |
|---|---|---|
| **IL10RB gRNA#1** | caccgAGGCTTGGCAGATGCACACG | aaacCGTGTGCATCTGCCAAGCCTc |
| **IL10RB gRNA#2** | caccgGGATCCTCGCAAGCTTTGAA | aaacTTCAAAGCTTGCGAGGATCCc |
| **IL10RB gRNA#3** | caccgGCATGCTGGAATGACGGTGG | aaacCCACCGTCATTCCAGCATGCc |
| **IL10RB gRNA#4** | caccgTTGAAGTCCGCTCTCCGCAC | aaacGTGCGGAGAGCGGACTTCAAc |
| **Scramble gRNA** | caccgGCACTCACATCGCTACATCA | aaacTGATGTAGCGATGTGAGTGCC |

**Supplementary Table 16: gRNA CRISPRa sequences**

### DESCRIPTION OF OTHER SUPPLEMENTARY APPENDICES

### SUPPLEMENTARY APPENDIX 2

**Brief description:** Table translating Phecodes to Phenotypes and the respective phenotype categories they belong to.

Column descriptions:

| Column | Description |
|---|---|
| Phecode | Numerical phecode |
| Phenotype | Phenotype associated with phecode |
| exclude_name | Category for each phecode. Phecodes were grouped into categories using Phecode Map v1.2 with manual curation for some uncategorized phecodes |

### SUPPLEMENTARY APPENDICES 3 TO 9

**Brief description:** TWAS results for COVID-19 GWASs:

| Name | Phenotype | TWAS results in |
|---|---|---|
| A1 | Very severe respiratory confirmed covid vs. not hospitalized covid | Supplementary Appendix 3 |
| A2 | Very severe respiratory confirmed COVID vs. population | Supplementary Appendix 4 |
| B1 | Hospitalized COVID vs. not hospitalized COVID | Supplementary Appendix 5 |
| B2 | Hospitalized COVID vs. population | Supplementary Appendix 6 |
| C1 | COVID vs. lab/self-reported negative | Supplementary Appendix 7 |
| C2 | COVID vs. population | Supplementary Appendix 8 |
| D1 | predicted COVID from self-reported symptoms vs. predicted or self-reported non-COVID | Supplementary Appendix 9 |

Each sheet within an excel file represents a different tissue. Column descriptions:

| Column | Description |
|---|---|
| gene | a gene's id: as listed in the Tissue Transcriptome model. Ensemble Id for most gene model releases (e.g. in this study). Can also be an intron's id for splicing model releases |
| gene_name | gene name as listed by the Transcriptome Model, typically HUGO for a |

| | |
|---|---|
| | gene (e.g. in this study). It can also be an intron's id. |
| zscore | S-PrediXcan or S-EpiXcan's association result for the gene |
| effect_size | S-PrediXcan or S-EpiXcan's association effect size for the gene. Can only be computed when beta from the GWAS is used. |
| pvalue | p-value of the aforementioned statistic |
| var_g | variance of the gene expression, calculated as W' * G * W (where W is the vector of SNP weights in a gene's model, W' is its transpose, and G is the covariance matrix) |
| pred_perf_r2 | $R^2_{CV}$ (cross-validated) of tissue model's correlation to gene's measured transcriptome (prediction performance). Recommended filtering is > 0.01 |
| pred_perf_pval | p-value of tissue model's correlation to gene's measured transcriptome (prediction performance). |
| pred_perf_qval | q-value of tissue model's correlation to gene's measured transcriptome (prediction performance). |
| n_snps_used | number of SNPs from GWAS that were used in the S-PrediXcan or S-EpiXcan analysis |
| n_snps_in_cov | number of SNPs in the covariance matrix |
| n_snps_in_model | number of SNPs in the imputation model |
| gwas | GWAS name (phenotype) from the COVID-19 hg |
| tissue | Imputation model used |
| method | Imputation method used for imputation model construction (in this study PrediXcan or EpiXcan) |
| gwas_fdr | FDR-adjusted p value [3] for association when considering all gene-trait associations across all tissue models within this specific GWAS (e.g. COVID-19 B2 phenotype) |
| Tissue.keyword | Pattern matching string for internal pipeline |
| fdr_all | FDR-adjusted p value [3] for association when considering all gene-trait associations across all tissue models and all GWASs (all COVID-19 phenotypes) |

| | |
|---|---|
| var_g | variance of the gene expression, calculated as W' * G * W (where W is the vector of SNP weights in a gene's model, W' is its transpose, and G is the covariance matrix) |

### SUPPLEMENTARY APPENDIX 10

**Brief description:** Results of *IL10RB* and *IFNAR2* GREx PheWAS

Column descriptions:

| Column | Description |
|---|---|
| ID | Concatenation of gene and phenotype analyzed |
| phecode | Phecode identifier in string format |
| beta | Association of gene expression and phenotype for gene and phenotype described in ID column |
| p | P value for association of gene expression and phenotype for gene and phenotype described in ID column |
| neg_log10p | $-\log_{10}$ transformation of p column |
| beta_dir | Binary classifier for direction of beta column. TRUE if positive. FALSE if negative. |
| beta_mag | Absolute value of beta column |
| phecode_num | Numerical phecode |
| Phenotype | Phenotype associated with each phecode |
| exclude_name | Category for each phecode. Phecodes were grouped into categories using Phecode Map v1.2 with manual curation for some uncategorized phecodes. Refer to Supplementary Appendix 2 for mappings. |
| HasCounts | Number of individuals in cohort with >0 counts for this phecode |
| NoCounts | Number of individuals in cohort with 0 counts for this phecode |
| gene | Gene whose expression was used in regression model for the specified phecode |
| adjusted.p | Adjusted p value using method specified in MC.method column |
| neg_log10adjusted.p | $-\log_{10}$ transformation of adjusted.p column |

| | |
|---|---|
| Rank | Rank of association significance from most significant to least |
| MC.method | Method for generating adjusted.p column from p column |

### SUPPLEMENTARY APPENDIX 11

**Brief description:** Computational drug repurposing results for launched compounds
First sheet: “Column.Info” provides the column descriptions for the results in the “launched.compounds sheet”.

### SUPPLEMENTARY APPENDIX 12

**Brief description:** Provides information about the EHR validation of the CDR pipeline.
Start from the first sheet of the Excel file (“TOC”)
